# From Text to Translation: Using Language Models to Prioritize Variants for Clinical Review

**DOI:** 10.1101/2024.12.31.24319792

**Authors:** Weijiang Li, Xiaomin Li, Ethan Lavallee, Alice Saparov, Marinka Zitnik, Christopher Cassa

## Abstract

**Backgrounds:** Despite rapid advances in genomic sequencing, most rare genetic variants remain insufficiently characterized for clinical use, limiting the potential of personalized medicine. When classifying whether a variant is pathogenic, clinical labs adhere to diagnostic guidelines that comprehensively evaluate many forms of evidence including case data, computational predictions, and functional screening. While a substantial amount of clinical evidence has been developed for many of these variants, the majority cannot be definitively classified as ‘pathogenic’ or ‘benign’, and thus persist as ‘Variants of Uncertain Significance’ (VUS).

**Methods:** We processed over 2.4 million plaintext variant summaries from ClinVar, employing sentence-level classification to remove content that does not contain evidence and removing uninformative or highly similar summaries. We then trained ClinVar-BERT to discern clinical evidence within these summaries by fine-tuning a BioBERT-based model with labeled records.

**Results:** We validated ClinVar-BERT model predictions for variant summaries that are classified as uncertain (VUS) using orthogonal functional screening data. ClinVar-BERT significantly separated estimates of functional impact in clinically actionable genes, including *BRCA1* (p = **1.90×10**^***−*20**^), *TP53* (p = **1.14×10**^***−*47**^), and *PTEN* (p = **3.82 × 10**^***−*7**^) and achieved an AUROC of 0.927 when classifying whether variants result in loss of function or have uncertain effects.

**Conclusion:** These findings suggest that ClinVar-BERT is capable of discerning evidence from diagnostic reports and can be useful for prioritizing variants for re-assessment by diagnostic laboratories and expert curation panels.

## Introduction

As genomic sequencing becomes increasingly integrated into clinical practice, the pace of variant interpretation and biomedical data production has accelerated. From 2019 to 2024, clinical laboratories have submitted 3.68 million variant classifications to ClinVar, a public archive of human genetic variation linked to clinical disorders 1. However, even in well-studied disease genes like *BRCA1* and *LDLR*, the majority of variants have only been observed in a few cases or a single individual 2, 3. Consequently, these variants often lack definitive human genetic evidence to be classified as pathogenic or benign and are instead classified as ‘Variants of Uncertain Significance’ (VUS) 4. Due to this clinical uncertainty, current practice guidelines do not recommend communicating information about VUS to providers or patients outside of a clinical indication for testing 5.

This translational gap prevents many patients who carry such variants in actionable genes from benefiting from genomic medicine at the population level 6. In nine genes that are responsible for hereditary breast and ovarian cancer (HBOC), Lynch syndrome (LS), and familial hypercholesterolemia (FH), there is a substantial burden of such rare variation in the population. Over 16% or 1 in 6 individuals carry a rare, non-synonymous variant, roughly 18-fold more than those already classified as pathogenic 7. Although many of these variants may have limited phenotypic effects, some carry a substantial risk of disease, but they lack sufficient evidence to be classified as pathogenic.

The sequence variant interpretation (SVI) process involves expert curation of evidence supporting pathogenicity or benignity, following guidelines developed by the American College of Medical Genetics and Genomics and the Association for Molecular Pathology (ACMG/AMP) 4. Information curated during assessment includes clinical case evidence, computational predictions of variant effect, experimental screening measuring protein function, among others. The available evidence is weighed collectively to reach a clinical classification for a variant and is often compiled into a text summary. Since 2019, 2.1 million of these variant submission text summaries have been deposited to ClinVar 1. Although they contain a great deal of curated evidence, these reports contain heterogeneous information, lack a consistent structure, and often do not use controlled vocabularies for evidence types or clinical information.

Understanding the evidence contained in these diagnostic reports can be useful for improving variant classification. Recent work has highlighted that as evidence of pathogenicity is developed for a variant, it can be used to sub-classify variants that may ultimately be classified as pathogenic 8. Here, we trained language models (LMs) to discern the evidence patterns that are indicative of pathogenicity, benignity, or uncertainty contained within variant summaries. Ultimately, this information can be used to identify VUS that have a substantial amount of evidence of pathogenicity to prioritize them for review by expert panels.

## Methods

We parsed and extracted 2.1 million plaintext submission summaries from ClinVar^1^. These submission summaries were generated by diagnostic labs to describe the evidence used during variant interpretation when submitting classifications to ClinVar 1. We processed these submission summaries by removing potential class labels from each submission summary record and filtering short or duplicated records, as described in the following section. This processing step reduced the number of submission summaries linked to variant assertions to about 1.2 million. We then developed training and testing datasets to fine-tune language models to understand evidence of pathogenicity. Next, we used these models to assign probabilities for each text summary for whether each variant is P/LP (pathogenic or likely pathogenic), B/LB (benign or likely benign), or VUS (variant of uncertain significance). Finally, we validated the classification accuracy of these models using text summary records as well as orthogonal functional screening data.

### ClinVar Dataset Processing

#### Removing short and duplicated text summaries

We filtered short submission summaries by setting a threshold of 100 characters to ensure each comment contained sufficient evidence and removed duplicate summaries from the dataset.

#### Removing highly similar submission summaries and standardizing text

Deduplication of text data has been shown to improve model performance significantly, especially for models with a large number of parameters, as this step removes redundancy and thereby increases the diversity of the data 9–12. Deduplication is frequently performed in an embedding space, where hashed numerical encodings are compared using methods such as MinHashLSH 13, which combines MinHash encoding 14 and the Locality Sensitive Hashing algorithm 15. Based on *n*-grams (a contiguous sequence of *n* characters/tokens from a given sample of text), MinHash provides a technique for quickly estimating the Jaccard similarity between two texts. In our study, we considered the sets of *n*-grams derived from the raw ClinVar text reports and studied similarity both across reports and within each report.

We performed deduplication both at the report level and the sentence level. Moreover, rather than relying on exact repetition searches, researchers have increasingly adopted “fuzzy” deduplication methods 11, 16, 17. This approach identifies “nearly” repeating data by measuring similarity and applying a threshold, and has been used in models such as GPT-3 18, Llama 19, Falcon 20, Pangu 21, and the Pile dataset 22. For our ClinVar model training, we incorporated fuzzy deduplication into our text preprocessing pipeline.

To standardize the text in the extracted ClinVar corpus, we took the following steps: First, we removed non-English words, including Unicode characters and Greek letters from the corpus. Next, we standardized punctuation and symbols to conform to conventional English usage. Finally, we applied MinHash 23 with a Jaccard similarity threshold of 95% to remove highly similar summaries based on groups of diagnostic labs and genes, as diagnostic labs may employ template-based summaries. Examples of deduplication results from ClinVar text are provided in Section Supplementary Methods.

#### SentenceClassifier

A ClinVar submission summary is a plaintext clinical report generated by a diagnostic lab that describes the evidence that was evaluated for whether a single variant may cause a specific phenotype or disorder. These submission text summaries often include one or more purely descriptive sentences, for example, describing the gene, location, and type of variant in the report, rather than the evidence that might be used to classify whether it is pathogenic or benign. Additionally, many text summaries include a conclusion sentence that summarizes the evidence described and an assertion about the variant’s pathogenicity, which maps to one of our three class labels (B/LB, VUS, and P/LP). Given our goal is to train the model to understand text representations of *evidence* that indicate that a variant is more likely to be pathogenic, benign, or uncertain, these conclusions could lead to bias or overfitting. To address this potential bias or overfitting, we fine-tuned a BERT 24 model with our labeled data to train a sentence classifier for identifying sentences in submission summaries as **description, evidence**, and **conclusion**. Examples of our labeled data are provided in Section Supplementary Methods.

We used the NLTK sentence tokenizer 25 to split each submission summary into individual sentences. We employed the SentenceClassifier to label sentences as description, evidence, and conclusion. We then removed description and conclusion sentences from each submission summary, only keeping evidence-labeled sentences. (see examples in Section Supplementary Methods). In this way, we could reduce the likelihood that our dataset contains assertions of variant pathogenicity. As a result of this text processing method, about 25% of the text is removed during this filtering process, detailed statistics on character, word, and sentence level are included in (b) in Figure 1a.

**Figure 1.**
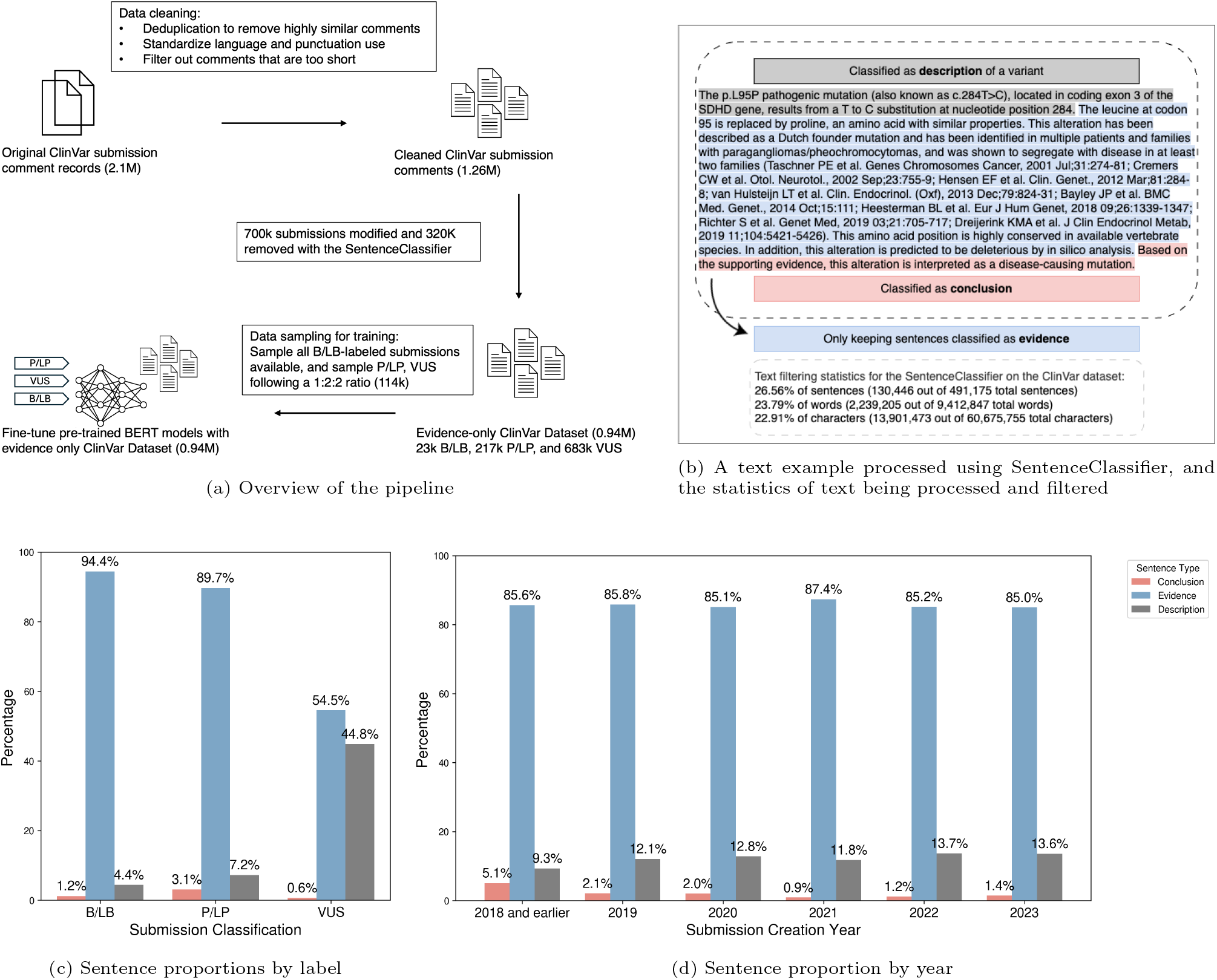
(a) An overview of text processing and record sampling used ahead of fine-tuning BERT models with ClinVar submission text summaries. (b) An example submission summary (SCV002749858): In this submission, the lab describes this variant (gray highlighting) and also classifies the variant as pathogenic (pink highlighting). We trained a sentence classifier to identify and filter these description and conclusion sentences so that only sentences containing evidence (blue highlighting) are used in model training. Additionally, we should the text filtering statistics for the SentenceClassifier on the ClinVar dataset. (c) Sentence type proportion distribution for three submission classification labels (B/LB, VUS, and P/LP) in the training data. Text summaries from the B/LB and P/LP classes have much larger fractions of evidence-labeled sentences, in contrast with VUS-labeled samples, which have a much larger share of description-labeled sentences. (d) Sentence type proportion distribution by ClinVar submission creation year, with pre-2019 years grouped together and individual years shown from 2019 through 2024. **Data source:** ClinVar data obtained from NCBI 1 (accessed in June 2023). Data and text corpus are processed using the pipeline discussed in the Methods Section.

#### Model training data processing

Finally, we sampled a subset of the ClinVar dataset to create training and testing datasets. Given the imbalanced distribution of B/LB submission summaries relative to P/LP and VUS summaries (22k B/LB, 216k P/LP, and 682k VUS), we sampled all B/LB summaries and constructed a 1:2:2 ratio for B/LB, P/LP, and VUS summaries. This corpus was split with 80% used for training and 20% for testing the model.

#### Fine-tuning BERT models for sequence classification

Upon obtaining training and testing data, we defined our approach as a sequence classification task. Specifically, we input a submission summary from ClinVar into a language model and task the model with predicting whether the variant is P/LP (pathogenic or likely pathogenic), B/LB (benign or likely benign), or VUS (variants of uncertain significance). We evaluated multiple BERT-based transformer models 26, including BERT 24, RoBERTa 27, BioBERT 28, ScholarBERT 29, and ClinicalBERT 30. BERT and RoBERTa are general-domain models, whereas BioBERT, ScholarBERT, and ClinicalBERT are domain-specific models pre-trained on large biomedical or clinical text corpora. Configurations and detailed training setups are discussed in Section Supplementary Methods.

### Model validation with experimental and computational data

#### Experimental and computational dataset construction

To validate the accuracy and generalizability of our fine-tuned models, we evaluated their performance on separately generated experimental screening data and computational predictions. For experimental screening data, we include estimates of functional impact for genetic variants derived from high-quality assays in genes with established clinical significance: *BRCA1, HRAS, LDLR, PTEN*, and *TP53*. Submission summaries for variants in these genes are excluded from the training and testing data. The DMS dataset was constructed by matching ClinVar submission summaries to variants with corresponding functional assay scores. These scores, downloaded from MaveDB 31, were processed using the FUSE optimization pipeline 32 and paired with ClinVar submission summaries at the amino acid substitution level. For validation using computational scores, REVEL scores and AlphaMissense scores are generated using Ensembl Variant Effect Predictor 33 for variants in the DMS dataset that intersect with ClinVar variants that have text summaries.

#### Benchmark evaluation using experimental functional scores

To evaluate the performance of our fine-tuned model using DMS data, we need to first generate ground truth labels. To do so, we assign labels (P/LP, B/LB, and VUS) based on functional scores. For functional data, there are three predicted impact types: ‘LOF’ or loss-of-function which is equivalent to our P/LP class label, ‘INT’ or intermediate, which is most equivalent to variants with our VUS class label, and ‘FUNC’ or functional, which is equivalent to our B/LB class label. Ground truth labels were applied to the functional datasets using two strategies: 1) Using the proportions of ClinVar VUS which are expected to truly be P/LP, VUS, or B/LB, which we derived from an early saturation genome editing study in *BRCA1* 34, by calculating the frequency of all ClinVar VUS at the time of publication which was found in the study to be LOF, INT, or FUNC. This resulted in assigning variant scores in the top 21.1 percentile as P/LP, variants below the 72.5 percentile as B/LB, and all remaining variants as VUS; 2) We also used an alternative approach with standardized threshold values to assign ground truth labels for each variant in the entire functional validation dataset. Variants in any gene that have a functional score of ≥ *μ* + *σ* are labeled as P/LP, functional scores that are *≤* 0 are labeled as B/LB, and the remaining variants are labeled VUS.

We calculated performance metrics as follows: We calculated pair-wise AUC values between each class (P/LP vs VUS, B/LB vs VUS, and P/LP vs B/LB) using ROC curve analysis, with the average AUC-ROC reported as the mean of these values. For overall performance metrics (accuracy, precision, recall, and F1 score), we used a class-weighted metrics calculation method to account for potential imbalances in our dataset. Results for strategy 1) for all models fine-tuned from text-processing methods are included in Table 2, and results for two label-assigning strategies are included in Table 3 in Section Supplementary Results.

**Table 1:** Performance of fine-tuned BioBERT-base models with ClinVar data using different text processing methods (evidence-only, rule-based filtered, and raw ClinVar data) compared to the pre-trained BioBERT-base model on the test data from the **raw-data** dataset.

| Model | Accuracy | Precision | Recall | F1 Score | AUC-ROC |
| --- | --- | --- | --- | --- | --- |
| BioBERT-base + ClinVar (evidence-only) | 0.9720 | 0.9721 | 0.9702 | 0.9711 | 0.9974 |
| BioBERT-base + ClinVar (rule-based) | 0.9805 | 0.9795 | 0.9802 | 0.9798 | 0.9987 |
| BioBERT-base + ClinVar (raw-data) | 0.7207 | 0.8157 | 0.7193 | 0.7264 | 0.9500 |
| BioBERT-base 28 | 0.3760 | 0.1601 | 0.3133 | 0.1919 | 0.5122 |

**Table 2:** Evaluation results of BioBERT-base models trained with three different text processing and sampling methods (evidence-only, rule-based filtered, and raw ClinVar data), compared to the pre-trained BioBERT-base baseline, evaluated on orthogonally generated DMS data. Ground truth labels are created using functional scores, with the 27.1 percentile as P/LP, 72.5 percentile as B/LB, and the rest being VUS.

| Model | Pair-wise AUC |  |  | Avg AUC-ROC | Accuracy | Precision | Recall | F1 Score |
| --- | --- | --- | --- | --- | --- | --- | --- | --- |
|  | P/LP vs B/LB | P/LP vs VUS | B/LB vs VUS |  |  |  |  |  |
| BioBERT-base + ClinVar (evidence-only) | **0.9272** | **0.8043** | 0.5470 | **0.7595** | 0.4753 | 0.4930 | 0.4753 | 0.4219 |
| BioBERT-base + ClinVar (rule-based) | 0.9096 | 0.7938 | 0.5377 | 0.7470 | 0.4891 | 0.5098 | 0.4891 | 0.4399 |
| BioBERT-base + ClinVar (raw-data) | 0.9037 | 0.7882 | 0.5826 | 0.7582 | 0.4840 | 0.5306 | 0.4840 | 0.4192 |
| BioBERT-base 28 | 0.3953 | 0.5428 | 0.3953 | 0.4503 | 0.2713 | 0.0736 | 0.2713 | 0.1158 |

**Table 3:** A comparison between ClinVar-BERT and BioBERT on embedding clustering quality of language model embeddings in the d-dimensional space. For each attribute, we report Silhouette scores (higher is better) and Davies-Bouldin indices (lower is better) as mean *±* standard deviation computed with 100 independent and randomly sampled runs.

| Attribute | ClinVar-BERT |  | BioBERT |  |
| --- | --- | --- | --- | --- |
|  | Silhouette | Davies-Bouldin | Silhouette | Davies-Bouldin |
| Clinical Significance | **0.4376 ± 0.0027** | **1.4352 ± 0.0091** | 0.0991 ± 0.0026 | 4.4854 ± 0.0555 |
| Submission Classification | **0.6446 ± 0.0020** | **0.5278 ± 0.0026** | 0.1170 ± 0.0029 | 3.2261 ± 0.0237 |
| Submitter | -0.2696 ± 0.0510 | 2.8505 ± 0.0883 | **0.1381 ± 0.0200** | **1.5892 ± 0.0453** |

#### Distribution analysis of model predictions with experimental and computational data

We then applied our fine-tuned models to make predictions on variants with submission summaries in the experimental and computational datasets, where the model would have three probability scores *P* (*B/LB*), *P* (*P/LP*), and *P* (*V US*) for each prediction. We then normalized prediction scores for B/LB and P/LP by 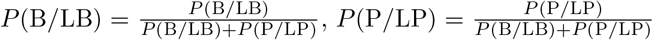 so that there is no overlap between the prediction scores for each group.

Then we recalibrated our predicted class label frequencies using the expected proportions of impacts from single nucleotide VUS from a well-established functional assay in *BRCA1* 34, employing the same approach as described above. Finally, with our recalibrated prediction labels, we performed Mann-Whitney U tests 35 to assess the statistical significance and median shifts between groups predicted as P/LP or B/LB based on their functional assay and computational scores. Results for ClinVar-BERT trained with evidence-only data are shown in Figure 2, Figure 3, and Figure 4. Results for all settings and models trained from different base models are shown in Figure 1, Figure 2, Figure 3, and Figure 4 in Supplementary Results Section (DMS validation results). Additionally, we evaluated our model against experimental functional scores using two alternative threshold approaches to ensure the robustness of our findings. First, we adopted the observed class distribution estimated from Figure 5 (B) in the AlphaMissense paper 36, where approximately 37% of MAVE prioritized variants are classified as P/LP and 49% as B/LB. Using these proportions, we labeled the top 37% of submissions by normalized prediction scores as P/LP and the bottom 49% as B/LB, with the remaining variants designated as VUS. Second, we sampled 10,000 VUS submissions from ClinVar across the five genes with available DMS functional scores *(BRCA1, TP53, LDLR, HRAS*, and *PTEN*) and applied the AlphaMissense-defined functional thresholds. This yielded a distribution of 61.26% B/LB, 30.57% P/LP, and 8.17% VUS, which we then used as thresholds for our model predictions. Results from these alternative threshold approaches are presented in Section Supplementary Results Figure 7 and Figure 8, demonstrating the consistency of our findings across different class distribution. Furthermore, we validated the model without applying recalibration, as shown in Figure 6. Overall, we validated the model with recalibration using three substantially different thresholds and without recalibration, and the results are consistent across all three settings.

**Figure 2.**
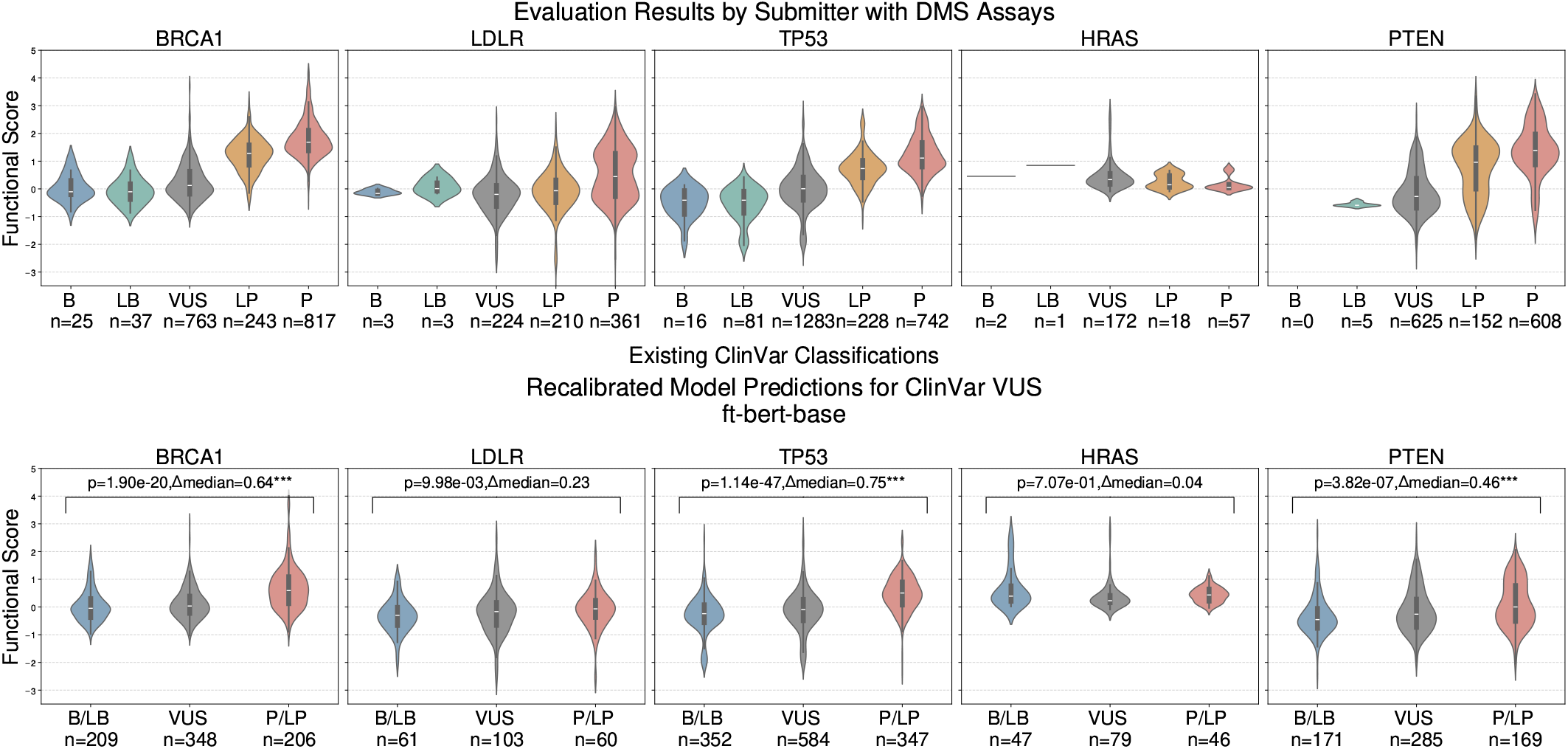
Top row: Existing classification on ClinVar (B, LB, VUS, LP, and P) on the *x*-axis and functional scores on the *y*-axis. Bottom row: recalibrated model prediction on **VUS**-labeled submission on the *x*-axis (B/LB, VUS, and P/LP), and functional scores on the *y*-axis. Statistical significance reported after Bonferroni correction (* *p<*0.05, ** *p<*0.01, *** *p<*0.001) **Data source:** The DMS dataset was constructed by matching ClinVar submission summaries to variants with corresponding functional assay scores. These scores, downloaded from MaveDB 31, were processed using the FUSE optimization pipeline 32 and paired with ClinVar submission summaries at the amino acid substitution level.

**Figure 3.**
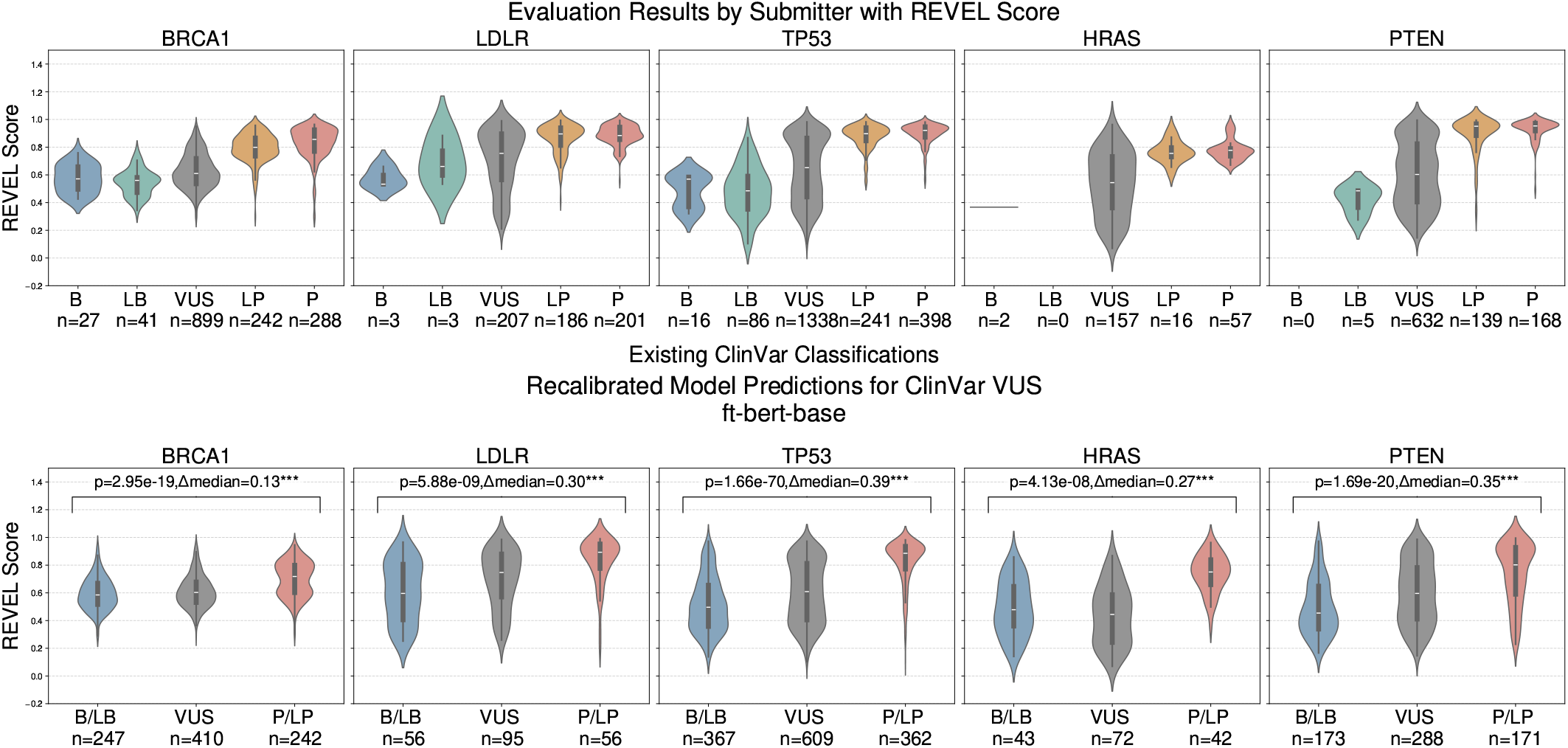
Top row: Existing classification on ClinVar (B, LB, VUS, LP, and P) on the *x*-axis and REVEL scores on the *y*-axis. Bottom row: recalibrated model prediction on **VUS**-labeled submission on the *x*-axis (B/LB, VUS, and P/LP), and REVEL scores on the *y*-axis. Statistical significance reported after Bonferroni correction (* *p<*0.05, ** *p<*0.01, *** *p<*0.001) **Data source:** REVEL score is generated using 33 using variant information for variants in the DMS dataset on ClinVar.

**Figure 4.**
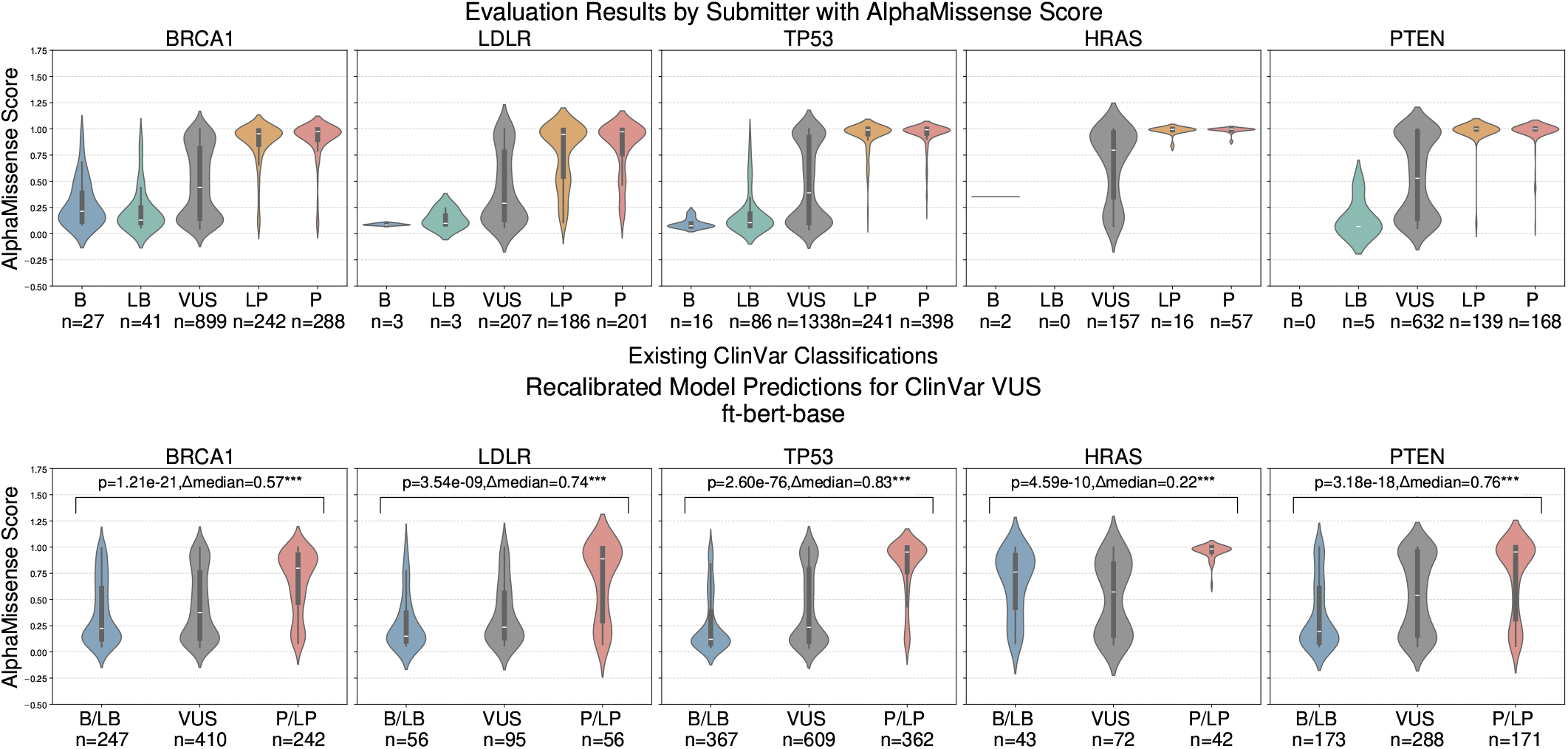
Top row: Existing classification on ClinVar (B, LB, VUS, LP, and P) on the *x*-axis and AlphaMis-sense scores on the *y*-axis. Bottom row: recalibrated model prediction on **VUS**-labeled submissions that are last updated **by 2023** on the *x*-axis (B/LB, VUS, and P/LP), and AlphaMissense scores on the *y*-axis. Statistical significance reported after Bonferroni correction (* *p<*0.05, ** *p<*0.01, *** *p<*0.001) **Data source:** AlphaMissense score is generated using 33 using variant information for variants in the DMS dataset on ClinVar.

**Figure 5.**
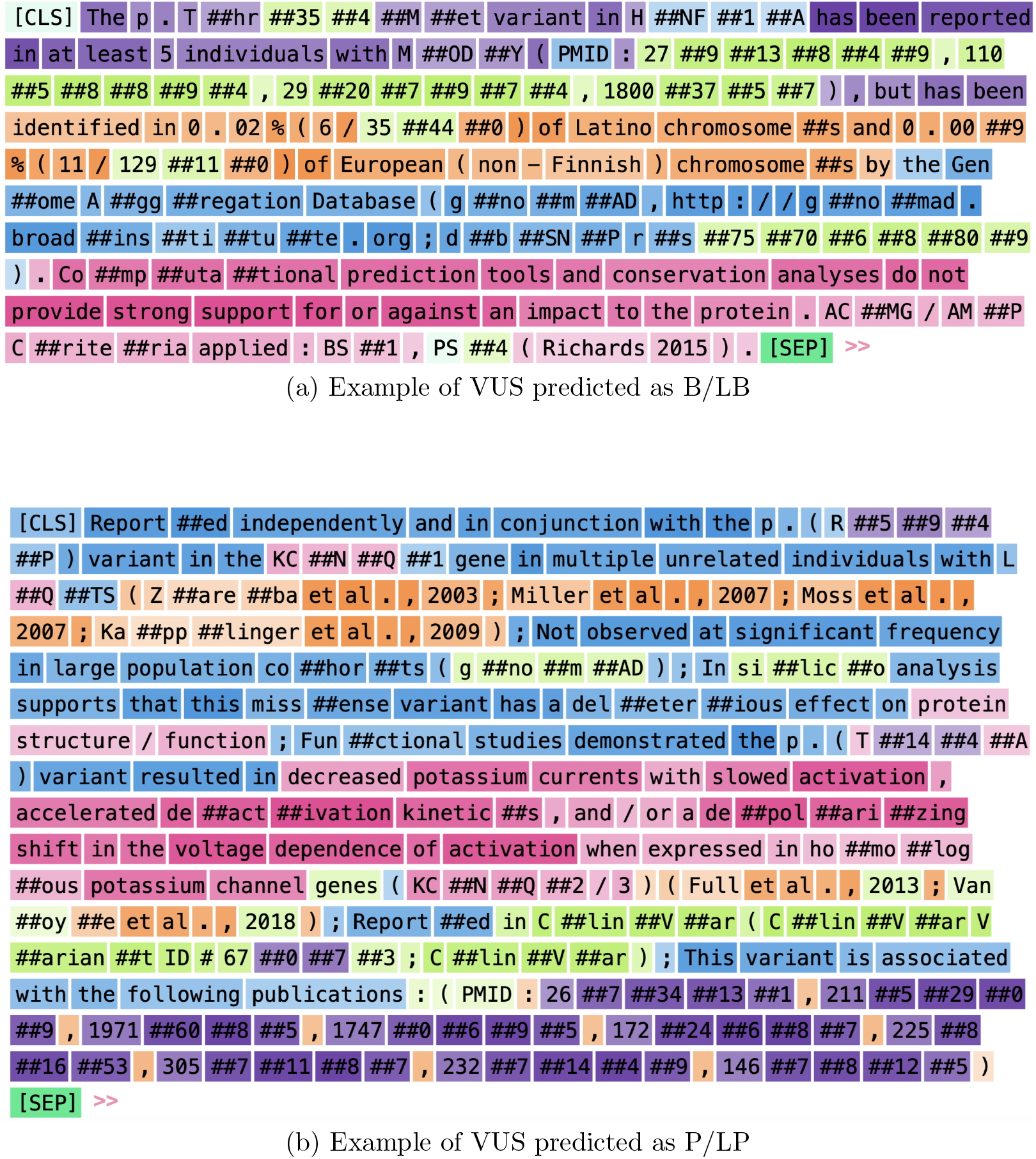
Neuron activation pattern visualization using Ecco for two variant classification case studies. Both examples are VUS submissions on ClinVar, but are predicted by our ClinVar-BERT model with high confidence (¿0.8) as B/LB and P/LP, respectively. (a) Example of a VUS predicted as B/LB, with strong neuron activations highlighting population frequency data (variant found in Latino and European chromosomes) and computational prediction results suggesting minimal impact on protein function. (b) Example of a VUS predicted as P/LP, with high neuron activations on sections describing functional evidence (decreased potassium currents, activation kinetics), case reports in unrelated individuals, and supporting literature citations, consistent with Long QT Syndrome pathogenicity. **Data source:** ClinVar data obtained from NCBI 1 (accessed in June 2023). We used our fine-tuned ClinVar-BERT to make predictions on the test data, and examples for making are obtained from the model prediction on the test data.

**Figure 6.**
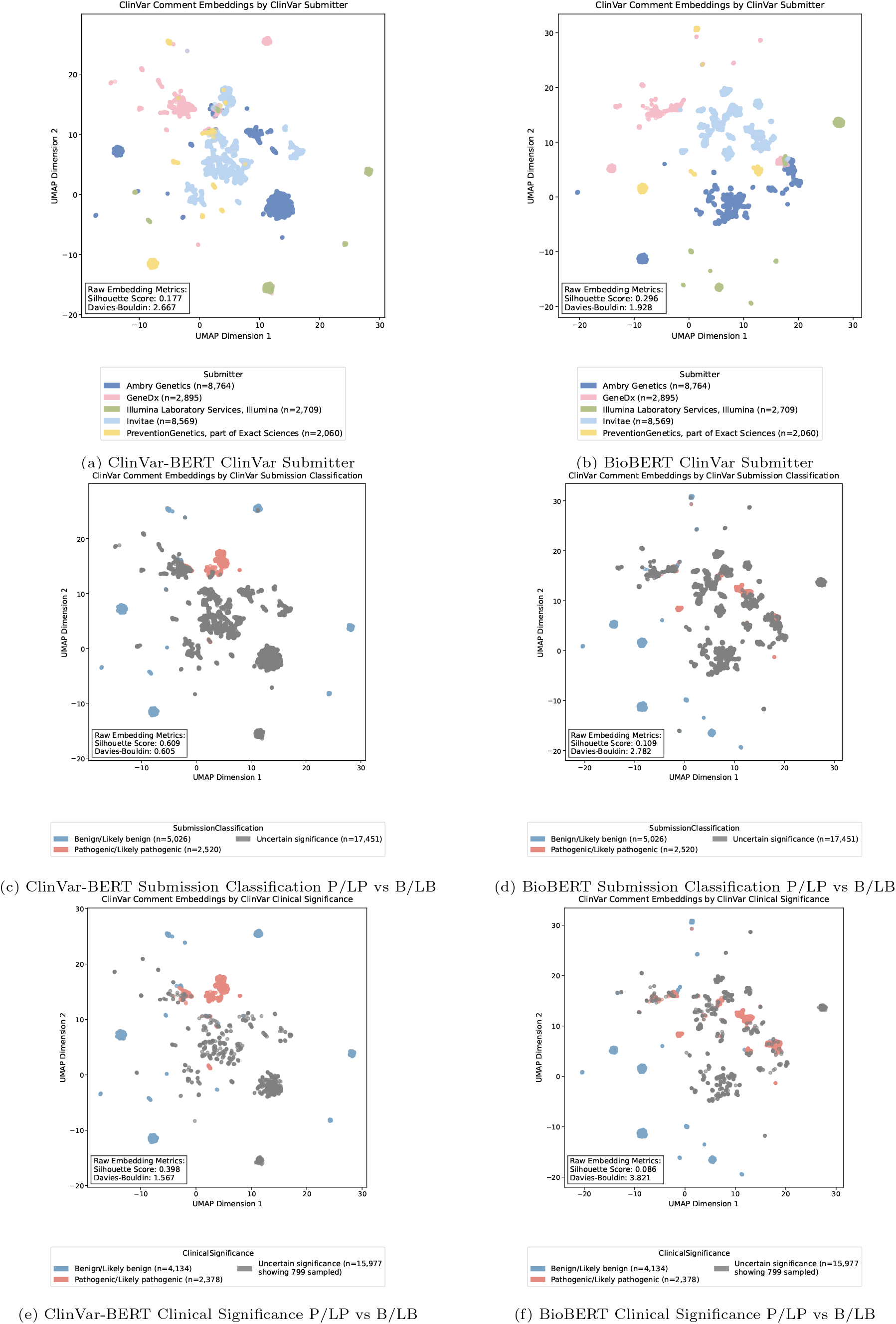
UMAP visualization comparing embeddings from ClinVar-BERT and BioBERT models for 25,000 randomly sampled ClinVar submission summaries. In each figure, we annotate clustering quality metrics computed with d-dimentional embedding directly from the model output. The left column visualizes ClinVar-BERT embeddings, and the right column visualizes BioBERT embeddings. (a) and (b) are visualizations with respect to the ClinVar submitter. (c) and (d) are UMAP visualizations colored with the submission classification, which is the variant pathogenicity assertion of the submission, showing P/LP and B/LB. (e) and (f) visualize the ClinVar clinical significance at the variant level on ClinVar, which considers all submissions related to the same variant and phenotype. **Data source:** ClinVar data obtained from NCBI 1 (accessed in June 2023). Embeddings for UMAP visualization are obtained by running a random sample of 25,000 text summaries through our fine-tuned ClinVar-BERT model. and the pre-trained BioBERT-base model 28.

### Neuron activation pattern visualization

Using the chosen preprocessing criteria and model, we generated predictions on the held-aside dataset. To analyze whether our ClinVar-BERT model focuses on words and phrases indicative of pathogenicity or benignity, we examined its neuron activation pattern. With the model tokenized ClinVar summaries, we utilized Ecco 37 to visualize and interpret neuron activation patterns. Ecco applies non-negative matrix factorization (NMF), a dimensionality reduction technique that transforms high-dimensional data into a lower-dimensional, more interpretable matrix. By specifying the number of components, we visualized the pattern of neuron activation through color-coded tokenizations. These components represent ‘concepts’ within the ClinVar summaries, highlighting relationships between words and phrases. This analysis revealed where the model focuses during the prediction task, providing insights into its interpretability.

#### Selection of summaries for attention weights visualization case review

Through a case review approach, we selected two ClinVar-labeled VUS summaries classified by the model as B/LB and P/LP respectively. To ensure strong case evidence, these summaries were sorted based on two criteria: high model prediction confidence and sufficient text length. We filtered summaries with a final class probability greater than 0.8 to ensure high model prediction confidence. Longer summaries were prioritized, as they are more likely to contain comprehensive textual evidence, with selection based on the total string length of the ClinVar summary. After sorting, we manually reviewed the summaries, focusing on those with evidence consistent with the American College of Medical Genetics and Genomics/Association of Molecular Pathology Sequence Variant Interpretation (ACMG/AMP SVI) framework 4.

### Submission summary embedding UMAP visualization

To visualize and compare the embeddings from the pre-trained BioBERT 28 model and our fine-tuned ClinVar-BERT model, we analyzed ClinVar submission summary text data focusing on their submitters and associated submission classification on ClinVar.

We first performed stratified sampling based on the ‘submitter’ to ensure a balanced representation while maintaining the natural distribution patterns in the data. Specifically, we selected the top 5 submitters by submission volume and sampled a total of 25,000 submissions, with a minimum of 1,000 samples per submitter. We computed embeddings for each submission summary in our sampled dataset using both BioBERT and ClinVar-BERT models, to conduct a comparative analysis of the embedding spaces before and after ClinVar-specific training. We then employed Uniform Manifold Approximation and Projection (UMAP) 38 for dimensionality reduction. The UMAP algorithm was configured with the following parameters: n neighbors=50 to balance local and global structure preservation, min dist=0.2, and cosine similarity as the distance metric to effectively capture semantic relationships between texts. The resulting two-dimensional embeddings were visualized as scatter plots, with points colored by three key categorical variables: ClinVar submitter, submission classification made by the submitter, and clinical significance (the interpretation of a variant).

## Results

Our objective is to train a model that learns text representations of evidence of variant pathogenicity, benignity, and uncertainty. With these learned representations of evidence, we then classify clinical text summaries for variants of uncertain significance (VUS), assessing how likely each variant is to contain evidence of being pathogenic, benign, or uncertain. A brief summary of the training strategy is provided in Figure 1a.

### Training data: Preprocessing variant text summaries

We first developed model training data using variants that had been previously classified by a clinical lab and deposited into ClinVar. A central challenge in learning representations of evidence is that ClinVar text summaries are heterogeneous with complex structures. We first deduplicated and filtered highly similar summaries, short, or uninformative summaries, and standardized punctuation, characters, and language (See Methods). We also sampled summaries across clinical labs, as many submissions come from just a few labs which could potentially contribute to bias and lack of text diversity in model training.

Text summaries from some clinical labs follow a template for how evidence is described. Consequently, summaries from those labs may exhibit high structural similarity. Furthermore, some sentences serve as a *conclusion* of the variant classification *(e*.*g. ‘Based on the supporting evidence, this alteration is interpreted as a disease-causing mutation*.*’)* which is a clear proxy for a class label. Other sentences provide a *description* of the variant *(e*.*g. ‘The p*.*L95P pathogenic mutation (also known as c*.*284T>C), located in coding exon 3 of the SDHD gene, results from a T to C substitution at nucleotide position 284*.*’)*. Both of these examples do not provide evidence of variant pathogenicity and could contribute to bias or overfitting in model training from the presence of specific structural elements.

We evaluated two approaches to mitigate the challenge of embedded class labels and structural similarity: using a rule-based method to filter these sentences and (2) using a sentence classifier to filter these sentences.

- **Rule-Based Filtering (‘rule-based’ dataset**): This approach uses a rule-based text processing pipeline to remove parts of sentences in the dataset that have pre-defined keywords and phrases that are suggestive of class labels, as specified in Supplementary Methods.
- **Sentence Classification Filtering (‘evidence-only’ dataset)**: This approach uses a model to classify sentences (SentenceClassifier) as *conclusion, description*, or *evidence*, and retains only evidence sentences. We classified and filtered predicted examples of these sentence types, as described in Figure 1b. Figure 1c describes the proportion of each type of sentence in the training data; Figure 1d shows the distribution of three sentence types by year.
- **Original Unfiltered Data (‘raw-data’ dataset)**: We contrast results with the original raw ClinVar data which has not been processed.

After filtering ClinVar text summaries using each of these three approaches, we created three distinct training sets. We sampled pathogenic or likely pathogenic (P/LP), uncertain (VUS), or benign or likely benign (B/LB), variants in proportions of 2:2:1, given the limited number of B/LB variants. All three sets contained the same number of variants, sampling was done by class, submitting lab, and gene.

### Evaluation of model performance

Next, we sought to evaluate the performance of each possible training set on *BioBERT-base*, a BERT-based model trained on a broad set of biomedical knowledge, including PubMed abstracts (PubMed) and PubMed Central full-text articles (PMC) 28. We fine-tuned *BioBERT-base* using each of our three training sets and measured classification performance using testing data, a random held-aside sample of 20% of the **raw-data** dataset without any text processing. We also compared to native *BioBERT-base* model performance without fine-tuning using ClinVar data. We observed a significant performance improvement for models fine-tuned with ClinVar data compared to the pre-trained *BioBERT-base* model. Results are shown in Table 1. This indicates that additional training with ClinVar data enhances the model’s ability to learn useful text representations and improves prediction accuracy on test data.

We observed significant evidence of overfitting for the three fine-tuned BioBERT base models we developed. Overfitting was most apparent with the model developed using the *raw-data* training set, in both classification performance and training loss. We found that training loss converged quickly for the *raw-data* approach, often within 100 steps. While the *rule-based* approach was slower to converge, both the *raw-data* and *rule-based* text processing methods converged quickly during training, achieving over 90% accuracy with fewer than 1,000 examples. In contrast, the model trained using the *evidence-only* approach was the slowest to converge. Training loss, evaluation loss, and accuracy during training for each text processing method can be found in Section Supplementary Methods (Figure 1c).

Next, we aimed to understand and mitigate potential causes of overfitting using an ablation study. For each variant summary, we identified the most influential sentence that could change model predictions (e.g., from P/LP to VUS, or *vice versa*), using randomly sampled ClinVar records. Detailed results of this study can be found in Section Supplementary Methods. We found that the model often learned to make predictions based on template-based structural characteristics, rather than learning unique or informative evidence of pathogenicity. We used this information to further refine the SentenceClassifier to help remove such template-based sentence structures to mitigate overfitting during training.

We additionally evaluated multiple pre-trained general-domain BERT language models, including BERT 24 and RoBERTa 27 on the **raw-data** test data. We also used BERT models trained with different biomedical text corpora including BioBERT 28, ClinicalBERT 30, and ScholarBERT 29 (See Methods), including base and large models where available, on the same test data from the **raw-data** dataset. Overall, pre-trained language models did not perform well in this ClinVar text classification task. There was still a noticeable performance difference between domain-specific language models and general-domain models, as shown in Table 2 in Section Supplementary Results.

Interestingly, we observed that the performance of domain-specific models, such as BioBERT and ScholarBERT, was not as strong as that of general-domain models, without further training on the ClinVar dataset. However, once these models were fine-tuned with ClinVar data, domain-specific models had slightly better performance than general-domain models, and the performance gap became smaller, with all fine-tuned models achieving comparable results.

### Validation using orthogonal functional screening data

We next proceeded to validate model predictions using orthogonal estimates of variant-level functional impact from deep mutational scanning (DMS) screens as ‘ground truth’ validation data. These experimental screens enable the functional assessment of thousands of coding variants that were installed in a cell line, typically replacing the native gene sequence. We evaluated model performance using DMS assays for five commonly screened genes: *BRCA1, LDLR, TP53, HRAS*, and *PTEN*, using functional score data from MaveDB 31, 39, which was normalized and processed using the FUSE pipeline 32.

We measured our model performance by comparing model predictions of pathogenicity from submission summaries with experimental measurements of variant functional impact from these DMS screening assays. We first developed class labels for ‘ground truth’ functional score data, using thresholds based on the prior expectation of proportions of single nucleotide ClinVar VUS 34. These thresholds were predicted to have damaging effects, intermediate effects, or have preserved function, from the *BRCA1* screening dataset using published threshold values. The resulting functional score thresholds set the top 27.1% of variant functional scores to be damaging (equivalent to our predicted P/LP class) and the bottom 27.5% of variant functional scores to have preserved function (equivalent to our B/LB class), with the rest labeled as having intermediate function (similar to some variants in our VUS class).

With each functional score having a ground truth class label, we then measured the accuracy of predictions from ClinVar-BERT to these ground truth labels. Results for models trained from different text processing methods and the base model are shown in Table 2. This shows that the model trained from the evidence-only dataset yields the best result, as it achieves the highest pair-wise AUC scores for P/LP vs B/LB and P/LP vs VUS. This demonstrates that this model can effectively separate P/LP from B/LB and VUS classes, indicating its potential uses in differentiating P/LP submissions from all submissions and could be useful for prioritizing submissions for further evaluation. We also evaluated a different method to label functional scores that are not based on any single gene, but rather on normalized scores for the entire functional validation dataset, and label variants with functional scores above +1*σ* as P/LP, variants with scores between 0 and 1*σ* for VUS, and variants with *<* 0*σ* as B/LB (see Methods). We find similar performance using this approach, and results for this additional method compared to the aforementioned threshold are shown in Table 3 in the Supplementary Results Section.

Additionally, we also evaluated ClinVar-BERT on different subsets of variants using DMS validation data, grouping by submitting laboratory, year range, and somatic or germline origin. We did not observe major differences in classification performance among different submitters or year ranges, but there are substantial differences for sample origin. There are considerably fewer submissions for variants of somatic origin, so this could potentially be useful to evaluate in future model development. Results are included in Table 5, Table 6, Table 7, and Table 8 in Section Supplementary Results (DMS validation results).

### Validation with experimental and computational data

In addition to benchmarking our fine-tuned models using labels derived from experimental functional scores, we also analyzed the distributions of functional scores and computational scores for model predictions on VUS submissions. We first analyzed DMS functional scores using existing ClinVar variant classifications (not derived from our models). Higher DMS functional scores generally indicate larger impacts on protein function, whereas lower scores indicate smaller impacts on protein function. As expected, in most genes, variants originally classified as B or LB had lower functional scores than VUS variants, which had lower functional scores than variants classified as LP or P (Figure 2, top). Notably, this was not the case for *HRAS* which had limited numbers of variants with classifications in ClinVar with functional scores, and *PTEN* which had no B variants and very few LB variants.

We then used our fine-tuned models to make predictions on each VUS-labeled ClinVar submission with a DMS functional score. Our models predicted that the large majority of VUS-labeled variants were VUS, which might limit the applicability of model predictions, so we sought to adjust VUS output class probabilities to be proportional to the true expected underlying proportions of B/LB, VUS, and P/LP classes. We evaluated several choices of prior expectation for these class proportions, and recalibrated model output class labels using these possible choices for evaluation. The first approach classified variant text summaries as B/LB, VUS, and P/LP following the proportion of the number of VUS that were found to retain function, to have intermediate effects, or to result in loss of function, using the number of ClinVar VUS at the time of publication, in an early comprehensive functional screening study 34 (See Methods). Following the same approach as discussed earlier, we also used computational scores to validate model performance, aware that there are potential limitations (see **??**). We evaluated the REVEL score 40 and AlphaMissense score 36 for validation, and we only consider submissions with the last update date by 2023 specifically for validation with AlphaMissense score (see Figure 2, Figure 3, and Figure 4). Results for models trained with different text processing methods are shown in Figure 1, and results for different base models are included in Section Supplementary Results (Figure 2, Figure 3, and Figure 4).

We also consider two alternative approaches to select these priors: 1) We adopted the observed class distributions estimated (37% of variants are classified as P/LP and 49% as B/LB.) from Figure 5 (B) in the AlphaMissense paper for variants that have been prioritized for MAVE screening 36; 2) We used an alternative distribution of 61.26% B/LB, 30.57% P/LP, and 8.17% VUS which we developed by sampling 10,000 VUS submissions from ClinVar across the five genes with available DMS functional scores (*BRCA1, TP53, LDLR, HRAS*, and *PTEN*) and applied AlphaMissense-defined score thresholds (See Methods). Results from these alternative threshold approaches are presented in Section Supplementary Results Figure 7 and Figure 8. Furthermore, we validated the model without applying any recalibration, as shown in Figure 6

The DMS functional scores of variants that were classified as P/LP were significantly different from the functional scores for variants that were predicted as B/LB, for nearly all model types and genes. For many models, we found that predicted P/LP and B/LB groups showed highly significant differences, with large separations in their median functional scores. Notably, DMS data from *LDLR* was only recently published in 41, so it was highly unlikely that this DMS information was present in any ClinVar text summaries that we evaluated for this gene. These results suggest that these fine-tuned models had been sufficiently trained to understand information useful for variant classification for variants within ClinVar that did not have sufficient evidence required to be classified as benign or pathogenic.

### Performance discussion

We found that the fine-tuned *BioBERT-base* and *BioBERT-large* models had the best performance across the language models we evaluated. We found consistent and highly significant differences between the functional scores of variants classified as B/LB and P/LP in all five genes, with large shifts in median functional scores between these two groups. Results for other fine-tuned models were included in Section Supplementary Methods (DMS validation results). For the remainder of the paper, we used the *evidence-only* trained version of *BioBERT-base*, which we call *ClinVar-BERT*.

### Neuron activation pattern visualization

Given that the types of evidence and descriptions in these submission summaries were heterogeneous, we aimed to characterize the forms of evidence that our models were identifying. We analyzed attention weights to understand the components of each text summary to identify the specific words or phrases that were influential in driving model classifications. We used Ecco 37, a Python library for interpreting and visualizing language model attention weights, to analyze neuron activation patterns within our ClinVar-BERT model when processing variant summaries. Ecco allows us to visualize how individual neurons in the model’s feed-forward neural network activate in response to specific input tokens. By tracking these activation patterns, we can identify which components of the text input or patterns contribute to the model’s predictions. This approach uses non-negative matrix factorization (NMF) to identify groups of neurons that are consistently being activated together in response to specific terms related to evidence used in variant interpretation summaries.

Using a case study approach, we analyzed examples of variants submitted to ClinVar as VUS that were classified by ClinVar-BERT as B/LB or P/LP with high model prediction confidence (probability of being P/LP or B/LB *>* 0.8). Figure 5 displays neuron activation patterns visualized using the NMF outputs from Ecco, where different colors represent groups of neurons that consistently co-activate in response to specific textual patterns.

While Ecco’s NMF outputs do not automatically label the concepts that are shown as different colors in the figure, the visualization reveals meaningful patterns in how our model processes variant evidence. We inspect each color separately and are able to identify distinct forms of clinical or experimental evidence that could be used in variant interpretation. In Figure 5(a), showing a VUS predicted as B/LB, we observe distinct neuron activation clusters (purple) highlighting case information (“has been reported in at least 5 individuals with MODY”); green-colored text shows publication identifiers, orange-colored text describing allele frequency data in different population groups, blue-colored text describing sources of population frequency data, and another cluster (pink) focusing on computational evidence (“prediction tools and conservation analyses do not provide strong support for or against an impact to the protein”).

Similarly, in Figure 5(b), illustrating a VUS submission predicted as P/LP by ClinVar-BERT, different neuron clusters activate in response to types of evidence patterns critical for variant interpretation. One cluster (blue) corresponds to case study evidence (“reported independently and in conjunction with,” “multiple unrelated individuals with”), another to functional evidence (colored in pink, “resulted in decreased potassium currents with slowed activation”), and a group in purple color is related to publication citations (“This variant is associated with the following publications”).

These activation patterns suggest that our model has learned to group related evidence types in a manner that aligns with evidence types that are used in the clinical variant interpretation process. The model appears to form internal representations that differentiate between different evidence types corresponding to the ACMG/AMP guidelines 4, such as variant population frequency data, computational scores, functional studies, and case reports. This behavior provides supporting evidence that ClinVar-BERT captures clinically meaningful text patterns during variant interpretation.

### Submission summary model embedding UMAP visualization

To investigate how fine-tuning with ClinVar evidence enhanced the ability of our language models to encode information potentially useful for variant interpretation, we performed a comparative analysis of model text embeddings for a set of submission summaries. We randomly sampled 25,000 ClinVar submission summaries and analyzed both the high-dimensional embedding space using clustering metrics and UMAP visualizations, for *BioBERT* 28 *and ClinVar-BERT*.

Our analysis finds substantial differences between BioBERT and ClinVar-BERT in how they represent ClinVar submission text summaries. As shown in Table 3, ClinVar-BERT produces significantly higher cluster separation for clinically relevant attributes compared to BioBERT. The Silhouette score, which measures how well samples are clustered by comparing the mean intra-cluster distance with the mean nearest-cluster distance (ranging from −1 to 1, with higher values indicating better-defined clusters), reveals important differences between ClinVar-BERT and BioBERT. In Table 3, we computed the average scores from 100 independent and random samples that we used the d-dimensional embeddings directly output from each language model. For clinical significance and submission classification, ClinVar-BERT’s raw embeddings achieve substantially higher Silhouette scores (0.4376 and 0.6446, respectively) compared to BioBERT (0.0991 and 0.1170). This difference is also reflected in the Davies-Bouldin (DB) index, a metric that quantifies the average similarity between each cluster and its most similar cluster, where lower values indicate better separation between clusters. ClinVar-BERT shows significantly lower DB index values (1.4352 and 0.5278) than BioBERT (4.4854 and 3.2261), indicating superior cluster separation, as also observed in ClinVar-BERT’s visualization in Figure 6e and Figure 6c, compared to BioBERT’s visualization as shown in Figure 6f and Figure 6d. However, this pattern is reversed for the Submitter attribute, where BioBERT shows better clustering (Silhouette score 0.1381) than ClinVar-BERT (−0.2696), as we could also observe in ClinVar-BERT’s UMAP visualization in Figure 6a compared to BioBERT’s Figure 6b. This suggests that fine-tuning may have shifted the model’s focus away from capturing submitter-specific patterns toward more variant interpretation-relevant features.

We also used UMAP to visualize the reduced embedding spaces of each variant grouping in Figure 6 (a)-(f). We note that dimensionality reduction through UMAP is primarily useful for ease of visualization of these high-dimensional embedding spaces, but the clustering metrics computed on the model output’s high-dimensional embeddings offer more reliable evidence of the models’ representational properties.

The distribution of VUS records of clinical significance (Figure 6e) from ClinVar-BERT embeddings reveals that the language patterns in VUS summaries systematically differ according to their proximity to P/LP versus B/LB clusters. VUS summaries clustering near P/LP regions tend to include some evidence of pathogenicity, as illustrated by this example: *“The G57R variant has not been published as pathogenic or been reported as benign to our knowledge. The G57R variant is not observed in large population cohorts (Lek et al*., *2016; 1000 Genomes Consortium et al*., *2015; Exome Variant Server)*…*This substitution occurs at a position that is conserved across species, and in silico analysis predicts this variant is probably damaging to the protein structure/function*.*”* This example contains population evidence (PM2) and computational evidence (PP3) of pathogenicity. Additionally, this variant has a REVEL score of 0.797 and an AlphaMissense score of 0.9931. Whereas VUS summaries that cluster closer to B/LB regions typically contain language suggesting an absence of pathogenic evidence or evidence of benignity, as seen in this example:*“In summary, the available evidence is currently insufficient to determine the role of this variant in disease*… *The threonine amino acid residue is found in multiple mammalian species, which suggests that this missense change does not adversely affect protein function*…*”*.

Moreover, we observed that VUS-variants that are closest to a P/LP cluster have multiple forms of evidence of pathogenicity. A series of detailed examples of ClinVar VUS submissions that are closest to a P/LP variant cluster are included in Table 5 in Section Supplementary Results (UMAP visualization clustering examples). We included computational evidence (REVEL and AlphaMissense score) where available, population frequency data from gnomAD, and variant functional consequences (e.g. missense, frameshift).

Overall, quantitative study via cluster quality metrics and qualitative analysis via UMAP visualization both suggest that fine-tuning with ClinVar data helped the model to adapt the embedding space to better capture text related to clinical evidence, and the underlying strength of that evidence. The model’s ability to capture the semantic features of clinical evidence can provide valuable insights for prioritizing variants for re-classification.

## Conclusion

In this study, we evaluated the potential of training language models to learn generalizable clinical evidence from unstructured variant summary text records in ClinVar. We fine-tuned both general-purpose and domain-specific models using labeled clinical summary text records to discern evidence of variant pathogenicity or benignity. We mitigated model bias and overfitting using a quality control pipeline that identified problematic records and a sentence classifier that filtered sentences that were unlikely to contain evidence of pathogenicity or benignity. We also validated our models using orthogonal data from functional screening assays and computational scores for genes that were entirely held aside from model training. We found that VUS classified as pathogenic and benign by *ClinVarBERT* had significantly different functional assay scores and computational scores, supporting the hypothesis that this model identified relevant evidence.

We found large improvements in variant classification tasks using models fine-tuned with ClinVar training data over general-domain models. Among the models we evaluated, classification performance was also consistently better when fine-tuning models pre-trained on broad biomedical text corpora, such as *BioBERT-base*, rather than using general-domain models. Given that *BioBERT-base* was trained on a large set of PubMed abstracts and PubMedCentral full-text articles, it appears to be a solid foundation for learning additional generalizable biomedical and clinical evidence within ClinVar text summaries. Training and classification metrics also demonstrate that our sentence classification approach (filtering sentences from text summaries that were unlikely to contain clinical evidence) performed well across validation datasets. This approach reduces overfitting, likely by removing conclusion sentences that contained proxy class labels, as well as removing structurally similar sentences, which were common among description text.

Our findings underscore the utility of language models in processing and interpreting intricate clinical narratives, offering potential applications in variant prioritization. Our model *ClinVar-BERT* has the potential to identify variants whose text summaries contain meaningful clinical evidence but were not yet sufficient for a pathogenic or benign classification. Clinical labs could choose to prioritize variants for reassessment that are close to meeting the evidence for a reclassification, and evaluate whether any new methods or sources of evidence that they consider in classification are now available that could update each variant classification. Similarly, ClinGen Variant Curation Expert Panels rely on biocurators and interpretation experts, so the ability to prioritize VUS with the strongest model predictions could help optimize which variants undergo a comprehensive evaluation by clinical experts.

Future research may include integrating information across a set of clinical summaries about the same variant from different diagnostic labs. By analyzing information developed by different labs, such an approach could prioritize variants that collectively had sufficient evidence to be reclassified, but where the evidence provided by any single lab was insufficient for classification. This information could also be used to inform ClinGen Variant Curation Expert Panels (VCEPs) about which variants are most likely to change classifications with expert review. Future work could also extract specific forms of evidence present within a clinical summary text, and use that to identify evidence gaps.

## Limitations of this work

This study has several limitations. First, some variants have multiple text summaries from different clinical labs, which may need to be harmonized. Because evidence of pathogenicity is often developed over time, more information is likely to be contained in the latest submission summary, but it may not include all available evidence. In contrast, while a very large number of variants have submission text summaries, many do not.

Second, text summaries are increasingly being generated using lab-standardized templates, which leads to high structural similarity among text summaries. This can lead to bias or overfitting if models learn template-specific features that are correlated with a specific classification, rather than learning about relevant evidence. Pre-processing approaches must meet the challenge of filtering or mitigating these highly predictive sentence structures. These templates are not uniform across labs, but increasingly VCEPs are recording structured evidence types with reports from expert reviews, which should help mitigate this issue more generally.

Furthermore, we acknowledge that there is potential bias and overfitting in our trained model that could affect the generalizability of performance. This could be attributable to multiple factors including the multi-stage training pipeline, syntactic similarity of text summaries, and previously available functional or computational studies. Moreover, there is a lack of independent test data. To address this as best as possible, we have used two forms of validation: functional assays and computational scores. Additionally, we also acknowledge there is potential circularity in using functional or computational scores for validation if that information might be included in submission summaries. We have attempted to mitigate this by evaluating a recently published functional dataset in our experimental validation, and by evaluating AlphaMissense scores restricted to variant summaries that were last updated prior to the publication of those scores. Notwithstanding, these scores are correlated with other existing computational estimates of pathogenicity, which could still potentially confound results.

Finally, while the accuracy of our classifications is strong, the most natural application of these models is for prioritizing variants for expert review. These models have not been calibrated to measure the strength of evidence provided for variant classification. Given that these models were trained on many forms of clinical evidence, it complicates their calibration and use when following the existing classification guidelines. Model performance is contingent on the quality and representativeness of the clinical reports available within ClinVar. Class imbalance, particularly in the B/LB category, poses a challenge that must continue to be addressed. Continual improvement of model architectures, training strategies, and dataset diversity is needed to enhance model robustness and generalizability.

In conclusion, by training language models to discern evidence of pathogenicity from unstructured clinical text, we have introduced a novel approach to prioritize variants for expert review. This research promises to allow clinicians to more readily make use of expert-curated information that is currently prohibitively complicated to use at scale. This information should enable expert panels to classify a larger proportion of variants as pathogenic or benign, allowing more patients to learn about clinically actionable findings. This could advance genomic medicine to the large number of patients who collectively harbor a VUS, potentially improving their clinical management.

## Data Availability

All data produced in the present study are available upon reasonable request to the authors

## Acknowledgments

We gratefully acknowledge funding from NIH R01HG010372 (W.L., E.L., C.C.) and R21HG014015 (W.L, M.Z., C.C.).

## Supplementary Methods

### Data cleaning and processing

With the dataset being parsed from the raw XML file, we then applied several text processing methods to prepare our dataset for model training and downstream evaluation tasks. Since ClinVar submission summary text is a specific domain of text such that it contains notations (i.e. HGVS nomenclature) and acronyms (i.e. ACMG evidence types) that only appear in this type of clinical text data, we develop text processing methods tailored to this dataset, the following describes our text processing steps in detail.

### Training SentenceClassifier

In order to remove variant or submission classification labels and their proxies in submission summaries via a robust approach, we train a sentence classifier for labeling a sentence as evidence or conclusion. This classifier aims to label sentences as evidence or conclusion. We define a sentence as conclusion if it represents a decision made by the submitter or the testing lab or institution, asserting a classification result for a specific case upon submission to ClinVar. References to classification results from other sources, such as other testing labs or publications, do not qualify as conclusion under this definition.

### Rule-based conclusion vs. evidence labeling

To construct a dataset for training the classifier, we initially implemented a rule-based labeling method, so that we could efficiently extract sentences labeled as conclusions from the ClinVar dataset. This rule-based methodology provides a more fine-grained approach to process data and is widely used in the field of machine learning and LLMs 42–45. In our case, our rules are various types of sentence patterns that we use for conclusion sentence matching. Analyzing the submission summaries from multiple submitters with various classification labels enables us to identify a unique list of keywords associated predominantly with conclusion sentences.

Using these keywords, we extracted a preliminary set of sentences, labeling them as conclusion. The remaining sentences were labeled as evidence. To enhance the reliability of this rule-based labeled dataset, we conducted manual reviews and corrections, resulting in a balanced dataset consisting of 2,500 evidence and 2,500 conclusion examples. Furthermore, to ensure data quality, a set of 100 randomly selected examples underwent a rigorous review by a domain expert.

#### Rule-Based Labeling: Conclusion Phrases and Keywords

- “In summary”
- “In conclusion”, “To summarize”, “To conclude”
- “Therefore,”, “is therefore predicted to be”
- “Taken together”, “Taking together”, “In brief”
- “this alteration remains unclear”, “Considering all the evidence”
- “Based on the available evidence”, “Due to insufficient evidences and the lack of functional studies”
- “After careful consideration”, “Upon review of the evidence”
- “Based on available information”, “Based on the results”
- “it has been classified as”, “Based on the available information”
- “based on the above information”, “with clinical assertions as classified by the original submitter”
- “Based on insufficient or conflicting evidence”, “the clinical significance of this alteration remains unclear”
- “Since supporting evidence”, “For these reasons”
- “based on the currently available information”, “We consider it to be”
- “As such”, “Due to these contrasting evidences and the lack of functional studies”
- “The score for this variant resulted in a classification of”, “Taking together”, “we classify this variant as”
- “we classify the”, “we classify it as”, “Considering that this is a”
- “there is insufficient evidence to classify”, “we interpret”
- “Considering available […]”
- “Due to limited information”, “Due to the potential impact of”
- “Based on the evidence outlined above”, “Variant of Uncertain Significance due to insufficient evidence:”
- “Since supporting an evidence is limited at this time”, “the clinical significance of this variant is”
- “Based on the classification scheme”, “Given all the evidence”
- “this collective evidence supports the classification of”, “leading us to conclude that”

### Examples of labeled conclusion vs evidence data

As we train a classifier for identifying sentences including classification labels in text, we first have a labeled dataset that includes balanced numbers of labeled examples for evidence and conclusion.

#### Example Labeled Data

**Conclusion-Labeled Data**

- The co-occurring 3’-UTR variant is located three base pairs upstream of the polyadenylation signal of PHEX, thus it remains unclear whether it is just a marker for this pathogenic duplication, or can be also detrimental in isolation.
- Therefore, this collective evidence supports the classification of the c.416G*>*A (p.Ser139Asn) as a recessive Likely Pathogenic variant for Nonsyndromic hearing loss and deafness.
- Thus, the clinical significance of the p.Phe17754Ser variant cannot be determined with certainty.
- Based on the collective evidence, the p.Arg947Pro variant is classified as a variant of uncertain significance for autosomal dominant pseudohypoaldosteronism type 1.
- In summary, the clinical signi ficance of the p.Arg343Gln variant is uncertain.
- Due to these contrasting evidences and the lack of functional studies, the clinical significance of the p.Glu886Ala change remains unknown at this time.

**Evidence-Labeled Data**

- This variant is present in population databases (rs201097255, gnomAD 0.06%).
- This population frequency is higher than expected for a pathogenic variant in MSH2 causing Lynch syndrome (BS1).
- ClinVar contains an entry for this variant (Variation ID: 17355).
- (I) 0304 - Variant is present in gnomAD (v2) *<*0.01 for a recessive condition (93 heterozygotes, 0 homozygotes).
- BARD1 His686Arg occurs at a position that is not conserved and is located in the BRCT 2 domain (UniProt).
- The K61E variant was not observed in approximately 6,500 individuals of European and African American ancestry in the NHLBI Exome Sequencing Project, indicating it is not a common benign variant in these populations.

### Examples of submission summaries before vs. after removing the conclusion-labeled sentence

#### Example summaries

**Before**

The c.1329delT pathogenic mutation, located in coding exon 5 of the BARD1 gene, results from a deletion of one nucleotide at nucleotide position 1329, causing a translational frameshift with a predicted alternate stop codon (p.V444Lfs*31). This alteration is expected to result in loss of function by premature protein truncation or nonsense-mediated mRNA decay. **As such, this alteration is interpreted as a disease-causing mutation**.

**After**

The c.1329delT pathogenic mutation, located in coding exon 5 of the BARD1 gene, results from a deletion of one nucleotide at nucleotide position 1329, causing a translational frameshift with a predicted alternate stop codon (p.V444Lfs*31). This alteration is expected to result in loss of function by premature protein truncation or nonsense-mediated mRNA decay.

With the sentence classifier, the conclusion sentence containing a proxy of the classification label “disease-causing mutation” is removed.

### Examples of highly similar submission summaries

With a threshold of **0.95**, the following submission summaries are identified as highly similar by MinHash 23

#### Example summaries

- This missense variant replaces methionine with isoleucine at codon 141 of the MSH2 protein. Computational prediction tool is inconclusive regarding the impact of this variant on protein structure and function. Internally defined REVEL score threshold: 0.5 *<* inconclusive *<* 0.7 (PMID: 27666373)…
- This missense variant replaces leucine with methionine at codon 9 of the MSH2 protein. Computational prediction is inconclusive regarding the impact of this variant on protein structure and function. Internally defined REVEL score threshold: 0.5 *<* inconclusive *<* 0.7 (PMID: 27666373)…
- This missense variant replaces methionine with valine at codon 779 of the MSH2 protein. Computational prediction tool is inconclusive regarding the impact of this variant on protein structure and function. Internally defined REVEL score threshold: 0.5 *<* inconclusive *<* 0.7 (PMID: 27666373)…
- This missense variant replaces lysine with glutamic acid at codon 579 of the MSH2 protein. Computational prediction is inconclusive regarding the impact of this variant on protein structure and function. Internally defined REVEL score threshold: 0.5 *<* inconclusive *<* 0.7 (PMID: 27666373)…

As we can see from the example above, with a threshold of 0.95 using MinHash, these submission summaries are identified as highly similar, and we can notice that these summaries are highly template-based and the only difference between each of them is the amino acid and codon information mentioned, and the rest of the evidence being reference to is exactly the same for all of them. These are the summaries that we want to filter out to ensure training text data diversity before sampling training and testing data.

### Fine-tuning language models with ClinVar dataset

#### Model training details

We fine-tuned all models using a maximum sequence length (max length) of 512 tokens, which is the maximum token length for BERT models. This configuration was chosen based on the distribution of token lengths in our training data, where the average token count is 155.59, the median is 143.0, and the 90th percentile is 254. We set the learning rate at 2 × 10^*−*5^ and applied a weight decay of 0.01 to optimize training.

Training loss, evaluation loss, and evaluation accuracy comparison during training using ClinVar datasets with different text processing methods:

**Fig. 1:**
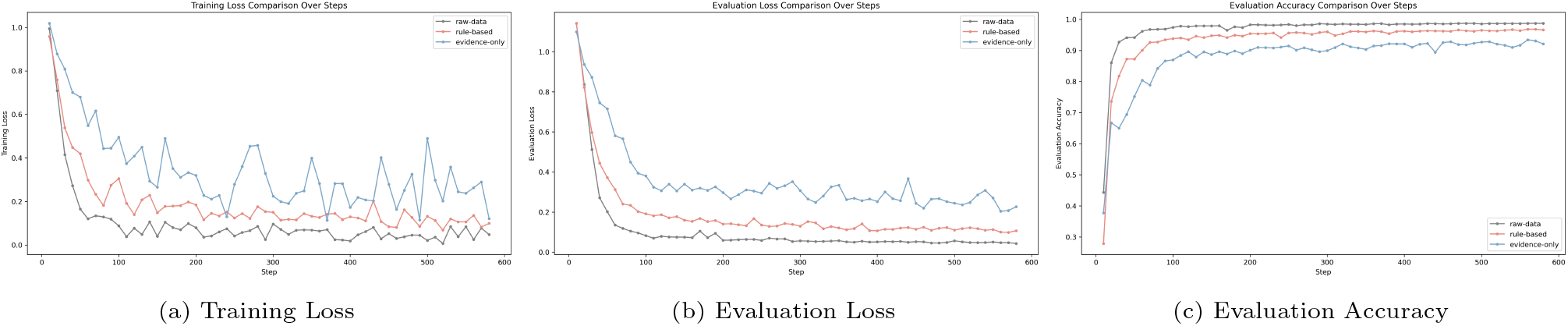
Comparison of training Loss, evaluation Loss, and accuracy during training among three text processing methods

#### ClinVar summary ablation study

##### Removing the sentence changes the prediction label from P/LP to VUS

~~~
{
       “Comment”: “This sequence change replaces arginine, which is basic
       and polar, with glutamine, which is neutral and polar, at codon 495
       of the MYBPC3 protein (p.Arg495Gln). This variant is present in
       population databases (rs200411226, gnomAD 0.006\%). This missense
       change has been observed in individuals with hypertrophic
       cardiomyopathy (PMID: 11499718, 20019025, 22857948, 23396983,
       24093860). ClinVar contains an entry for this variant (Variation ID:
       164113). Algorithms developed to predict the effect of missense
       changes on protein structure and function (SIFT, PolyPhen-2, Align
       GVGD) all suggest that this variant is likely to be disruptive. This
       variant disrupts the p.Arg495 amino acid residue in MYBPC3. Other
       variant(s) that disrupt this residue have been determined to be
       pathogenic (PMID: 18403758, 19659763, 20624503). This suggests that
       this residue is clinically significant, and that variants that
       disrupt this residue are likely to be disease-causing.”,
       “removed_sentence”: “This missense change has been observed in
       individuals with hypertrophic cardiomyopathy (PMID: 11499718,
       20019025, 22857948, 23396983, 24093860).”,
       “Original_Prediction”: “P/LP”,
       “Ablated_Prediction”: “VUS”,
       “Prediction_Difference”: “Changed”,
       “SCV”: “SCV000218744”,
       “Submitter”: “Invitae”,
       “Gene”: “MYBPC3”,
       “ground_truth_classification”: “P/LP”,
       “prediction_labels_ft”: “P/LP”,
       “prediction_scores_ft”: [
             0.99008584,
             0.009653065,
             0.00026108805
      ],
       “Ablated_Prediction_Scores”: [
             8.087950845947489e-05,
             0.9998732805252075,
             4.587376315612346e-05
      ],
       “Score_Difference”: 0.9900049604915405
}
~~~

##### VUS to P/LP Influential Sentences ExamplesUS to P/LP Influential Sentences Examples

- This sequence change creates a premature translational stop signal (p.Arg494*) in the EGF gene.
- This sequence change disrupts the translational stop signal of the FANCM mRNA.
- This sequence change affects an acceptor splice site in intron 5 of the TBX20 gene.
- The V462I variant in the PCCB gene has not been reported previously as a pathogenic variant, nor as a benign variant, to our knowledge.
- This variant results in a copy number gain of the genomic region encompassing exon(s) 50-57 of the NF1 gene.

##### VUS to B/LB Influential Sentences Examples

- This sequence change replaces aspartic acid, which is acidic and polar, with glycine, which is neutral and non-polar, at codon 650 of the BBS9 protein (p.Asp650Gly).
- This sequence change replaces arginine, which is basic and polar, with threonine, which is neutral and polar, at codon 429 of the PIGV protein (p.Arg429Thr).
- This sequence change replaces aspartic acid, which is acidic and polar, with glycine, which is neutral and non-polar, at codon 435 of the SMCHD1 protein (p.Asp435Gly).
- This sequence change replaces arginine, which is basic and polar, with cysteine, which is neutral and slightly polar, at codon 261 of the DHX32 protein (p.Arg261Cys).
- This sequence change replaces threonine, which is neutral and polar, with proline, which is neutral and non-polar, at codon 249 of the DSC2 protein (p.Thr249Pro).

### Analysis

#### Neuron activation pattern analysis

The Ecco visualization model is configured by specifying the model id, activations, and model config. For model id, the path to the trained model was provided, and activations were enabled (True). The model config was defined with several parameters: embedding set to ‘embeddings.word embeddings’, type as ‘mlm’, activations as ‘intermediate textbackslash.dense’, token prefix as ‘, ‘, and partial token prefix as ‘##’.

## Supplementary Results

### Distribution of sentence type across ClinVar dataset

**Table 1:**
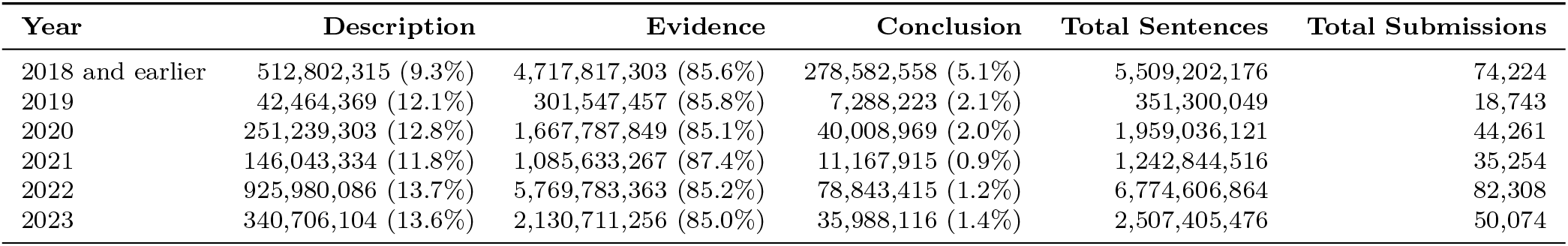
Distribution of type of sentences across the ClinVar dataset grouped by the year of ClinVar submission creation.

| Year | Description | Evidence | Conclusion | Total Sentences | Total Submissions |
| --- | --- | --- | --- | --- | --- |
| 2018 and earlier | 512,802,315 (9.3%) | 4,717,817,303 (85.6%) | 278,582,558 (5.1%) | 5,509,202,176 | 74,224 |
| 2019 | 42,464,369 (12.1%) | 301,547,457 (85.8%) | 7,288,223 (2.1%) | 351,300,049 | 18,743 |
| 2020 | 251,239,303 (12.8%) | 1,667,787,849 (85.1%) | 40,008,969 (2.0%) | 1,959,036,121 | 44,261 |
| 2021 | 146,043,334 (11.8%) | 1,085,633,267 (87.4%) | 11,167,915 (0.9%) | 1,242,844,516 | 35,254 |
| 2022 | 925,980,086 (13.7%) | 5,769,783,363 (85.2%) | 78,843,415 (1.2%) | 6,774,606,864 | 82,308 |
| 2023 | 340,706,104 (13.6%) | 2,130,711,256 (85.0%) | 35,988,116 (1.4%) | 2,507,405,476 | 50,074 |

### Test data evaluation results

Results on the test data for fine-tuned model using *evidence-only* training data and pre-trained model.

**Table 2:** Performances of fine-tuned BERT models using evidence-only training data compared to pre-trained models on ClinVar raw-data test data.

| Model | Accuracy | Precision | Recall | F1 Score | AUC-ROC |
| --- | --- | --- | --- | --- | --- |
| BioBERT-large 28 + ClinVar | 0.9754 | 0.9756 | 0.9730 | 0.9743 | 0.9982 |
| ScholarBERT 29 + ClinVar | 0.9720 | 0.9721 | 0.9702 | 0.9711 | 0.9974 |
| ClinicalBERT 30 + ClinVar | 0.9666 | 0.9685 | 0.9621 | 0.9651 | 0.9976 |
| RoBERTa-base 27 + ClinVar | 0.9717 | 0.9734 | 0.9669 | 0.9700 | 0.9979 |
| RoBERTa-large 27+ ClinVar | 0.9729 | 0.9728 | 0.9705 | 0.9716 | 0.9978 |
| BERT-Base 24 + ClinVar | 0.9729 | 0.9724 | 0.9715 | 0.9720 | 0.9979 |
| BERT-Large 24 + ClinVar | 0.9754 | 0.9756 | 0.9730 | 0.9743 | 0.9982 |
| BioBERT-large | 0.2000 | 0.3046 | 0.3318 | 0.1136 | 0.5599 |
| ScholarBERT | 0.2309 | 0.3277 | 0.3477 | 0.1789 | 0.5344 |
| ClinicalBERT | 0.4152 | 0.2801 | 0.3505 | 0.2346 | 0.4686 |
| RoBERTa-base | 0.1360 | 0.0900 | 0.1537 | 0.1110 | 0.4988 |
| RoBERTa-large | 0.3030 | 0.2325 | 0.3774 | 0.2552 | 0.5984 |
| BERT-base | 0.4025 | 0.4503 | 0.3355 | 0.1953 | 0.3768 |
| BERT-large | 0.1998 | 0.2139 | 0.3132 | 0.1372 | 0.4705 |

### DMS validation results

**Table 3:**
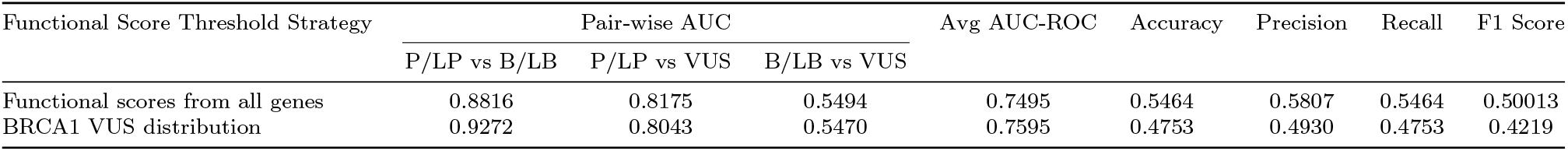
Evaluation results of evidence-only ClinVar-BERT model, using different thresholds for generating ground truth labels for functional scores. We label each variant in the validation set using threshold values based on their functional scores: First row: Variants in any gene that have a functional score of *≥ µ* + *σ* are labeled as P/LP, functional scores that are *≤* 0 are labeled as B/LB, and the remaining variants are labeled VUS; Second row: We measured the proportions of ClinVar VUS at time of publication for a large saturation genome editing screen in BRCA1, identifying how many are functional, indeterminate, or loss-of-function using published functional screening threshold values. This results in a variant labeling strategy that assigns functional scores that are *≥* 72.9 percentile as P/LP, *≤* 27.5 percentile labeled as B/LB, and the remaining are labeled as VUS.

**Table 4:**
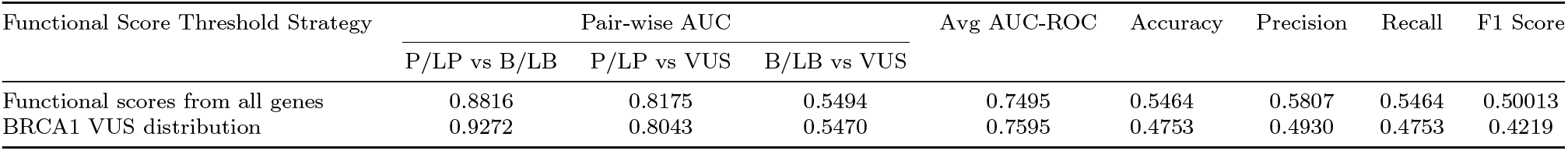
Evaluation results of evidence-only ClinVar-BERT model, using different thresholds for generating ground truth labels for functional scores. We label each variant in the validation set using threshold values based on their functional scores: First row: Variants in any gene that have a functional score of *≥ µ* + *σ* are labeled as P/LP, functional scores that are *≤* 0 are labeled as B/LB, and the remaining variants are labeled VUS; Second row: We measured the proportions of ClinVar VUS at time of publication for a large saturation genome editing screen in BRCA1, identifying how many are functional, indeterminate, or loss-of-function using published functional screening threshold values. This results in a variant labeling strategy that assigns functional scores that are *≥* 72.9 percentile as P/LP, *≤* 27.5 percentile labeled as B/LB, and the remaining are labeled as VUS.

**Table 5:**
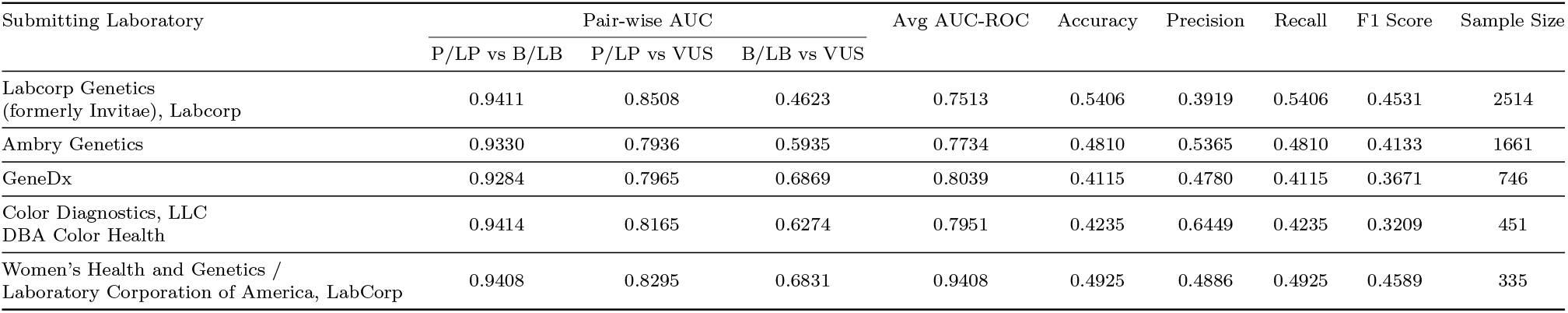
Performance of evidence-only ClinVar-BERT evaluated using functional score–based ground truth labels derived from BRCA1. Variants are labeled as P/LP if their functional scores fall within the top 27.1%, B/LB if within the bottom 27.5%, and VUS otherwise. The table reports the model’s classification performance across submissions from different laboratories.

**Table 6:** Performance of evidence-only ClinVar-BERT evaluated using functional score–based ground truth labels derived from BRCA1. Variants are labeled as P/LP if their functional scores fall within the top 27.1%, B/LB if within the bottom 27.5%, and VUS otherwise. The table reports the model’s classification performance across submissions from ranges of years.

| Year Range | Pair-wise AUC |  |  | Avg AUC-ROC | Accuracy | Precision | Recall | F1 Score | Sample Size |
| --- | --- | --- | --- | --- | --- | --- | --- | --- | --- |
|  | P/LP vs B/LB | P/LP vs VUS | B/LB vs VUS |  |  |  |  |  |  |
| 2022 - 2024 | 0.9293 | 0.8182 | 0.5558 | 0.7678 | 0.4552 | 0.5278 | 0.4552 | 0.3877 | 2478 |
| 2019 - 2021 | 0.9254 | 0.8121 | 0.5949 | 0.7775 | 0.5260 | 0.5216 | 0.5260 | 0.4769 | 2152 |
| Before 2019 | 0.8964 | 0.7807 | 0.5778 | 0.7516 | 0.5037 | 0.4869 | 0.5037 | 0.4585 | 1336 |

**Table 7:** Performance of evidence-only ClinVar-BERT evaluated using functional score–based ground truth labels derived from BRCA1. Variants are labeled as P/LP if their functional scores fall within the top 27.1%, B/LB if within the bottom 27.5%, and VUS otherwise. The table reports the model’s classification performance across submissions from different variant origins using submissions last updated by 2023.

| Variant Origin | Pair-wise AUC |  |  | Avg AUC-ROC | Accuracy | Precision | Recall | F1 Score | Sample Size |
| --- | --- | --- | --- | --- | --- | --- | --- | --- | --- |
|  | P/LP vs B/LB | P/LP vs VUS | B/LB vs VUS |  |  |  |  |  |  |
| Germline | 0.9282 | 0.8295 | 0.5592 | 0.7723 | 0.5122 | 0.5112 | 0.5122 | 0.4519 | 5107 |
| Germline/Somatic | 0.7954 | 0.6935 | 0.5786 | 0.6891 | 0.3690 | 0.6176 | 0.3690 | 0.3436 | 767 |
| Not Provided | 0.7437 | 0.5517 | 0.5053 | 0.6002 | 0.3415 | 0.2454 | 0.3415 | 0.2242 | 82 |

**Table 8:** Performance of evidence-only ClinVar-BERT evaluated using functional score–based ground truth labels derived from BRCA1. Variants are labeled as P/LP if their functional scores fall within the top 27.1%, B/LB if within the bottom 27.5%, and VUS otherwise. The table reports the model’s classification performance across submissions from the different variant origins for the PTEN gene using submissions last updated by 2023.

| Variant Origin<br>for PTEN Gene | Pair-wise AUC |  |  | Avg AUC-ROC | Accuracy | Precision | Recall | F1 Score | Sample Size |
| --- | --- | --- | --- | --- | --- | --- | --- | --- | --- |
|  | P/LP vs B/LB | P/LP vs VUS | B/LB vs VUS |  |  |  |  |  |  |
| Germline | 0.9230 | 0.7247 | 0.5331 | 0.7269 | 0.4561 | 0.5927 | 0.4561 | 0.3720 | 1151 |
| Germline/Somatic | 0.8031 | 0.6212 | 0.4800 | 0.6348 | 0.3214 | 0.2139 | 0.3214 | 0.2152 | 84 |

**Table 9:**
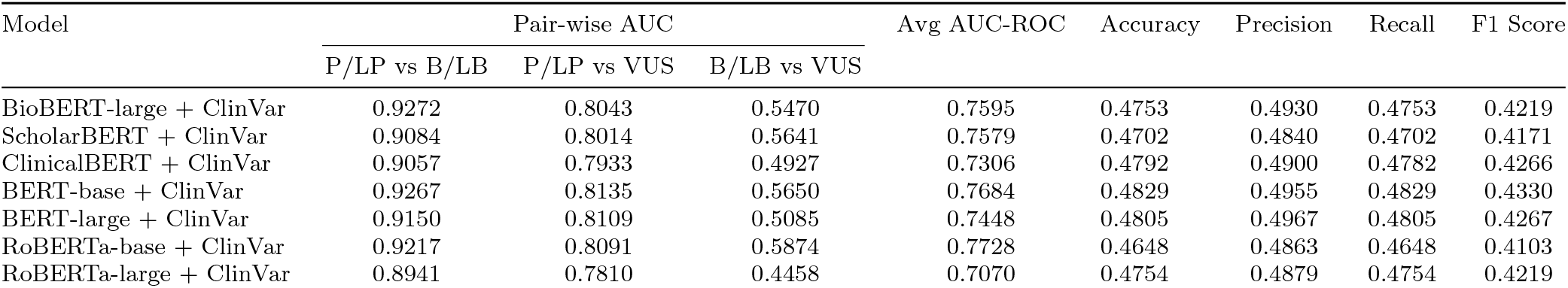
Evaluation results of fine-tuned models trained with the evidence-only dataset on orthogonally generated DMS data. Variants are labeled as P/LP if their functional scores fall within the top 27.1%, B/LB if within the bottom 27.5%, and VUS otherwise.

**Fig. 1:**
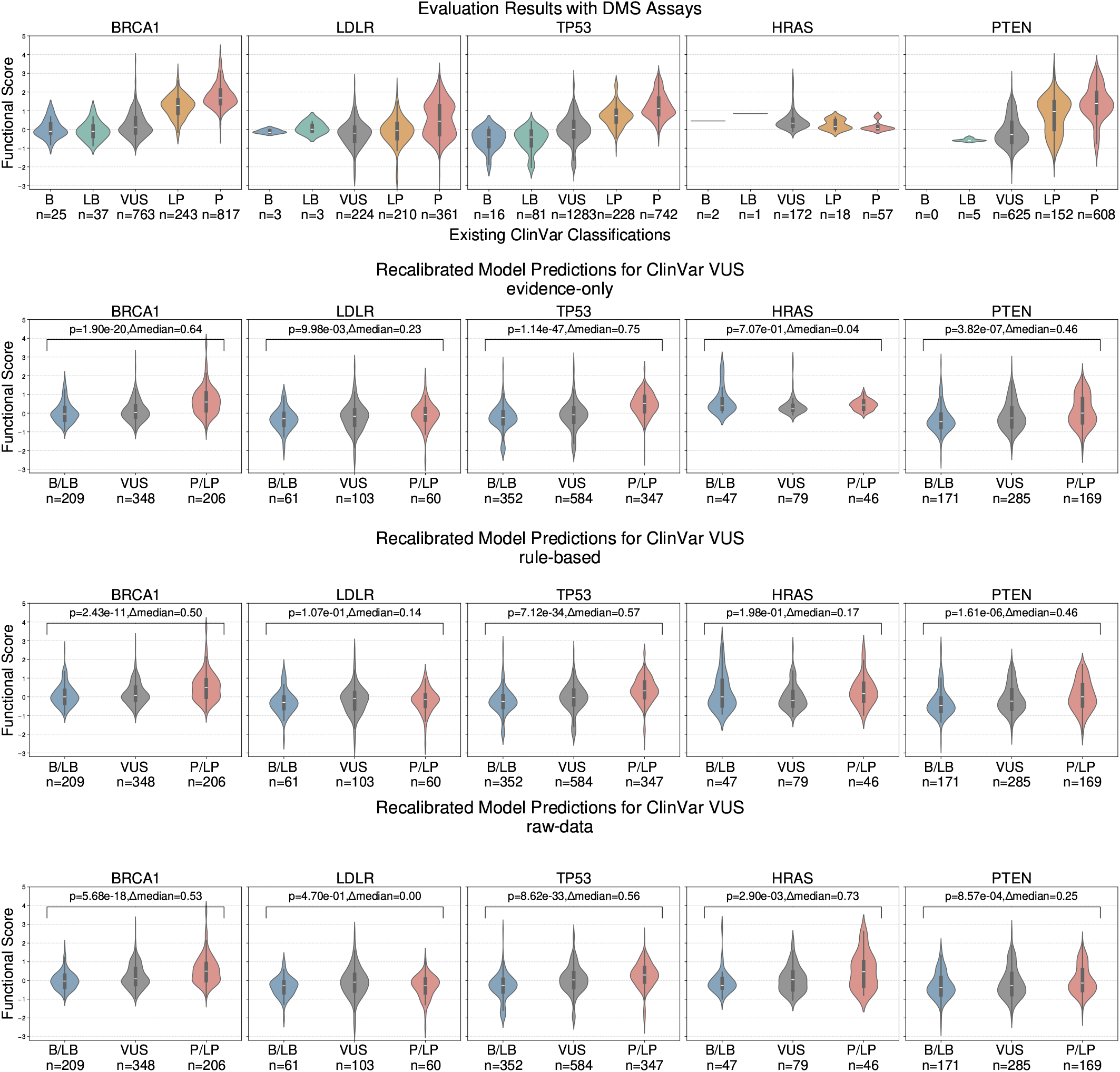
Top row: Existing classification on ClinVar (B, LB, VUS, LP, and P) on the *x*-axis and functional scores on the *y*-axis. The following rows are recalibrated model prediction on **VUS**-labeled submission on the *x*-axis (B/LB, VUS, and P/LP), and functional scores on the *y*-axis.

**Fig. 2:**
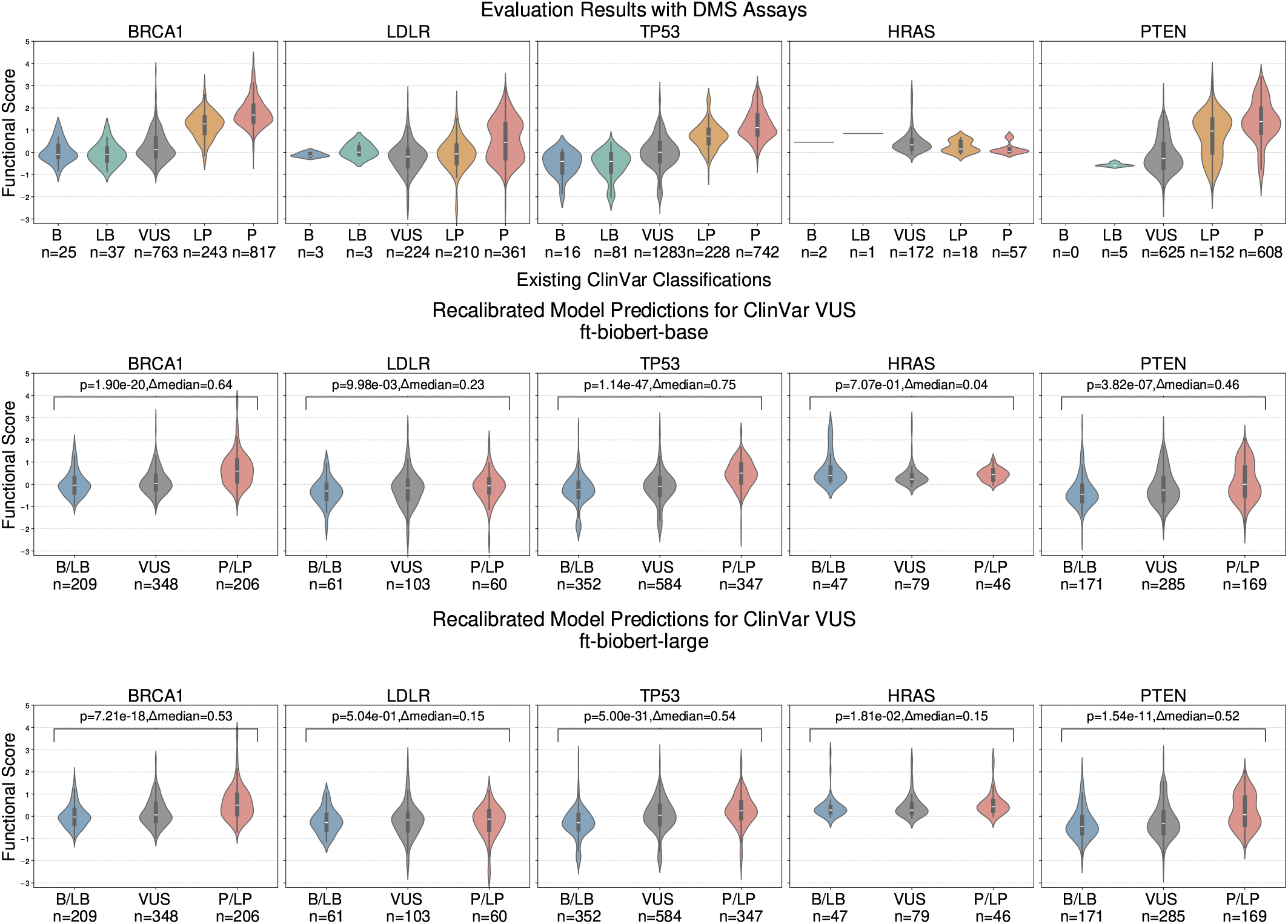
Comparison of DMS validation results of **evidence-only** trained BioBERT-base and BioBERT-large models on orthogonally generated DMS data

**Fig. 3:**
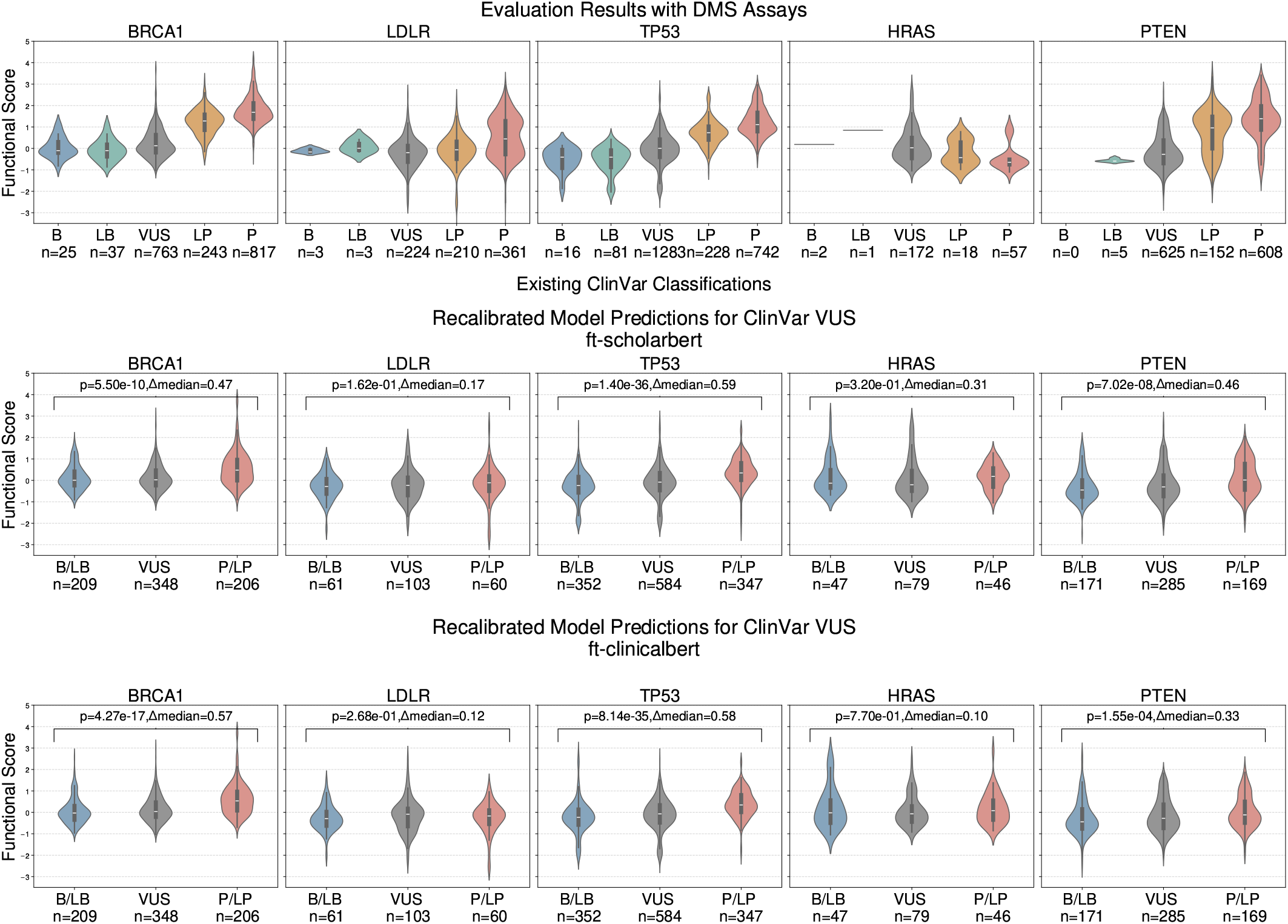
A comparison of two domain-specific BERT-architecture models (ScholarBERT and ClinicalBERT) fine-tuned with **evidence-only** training data performances on DMS validation.

**Fig. 4:**
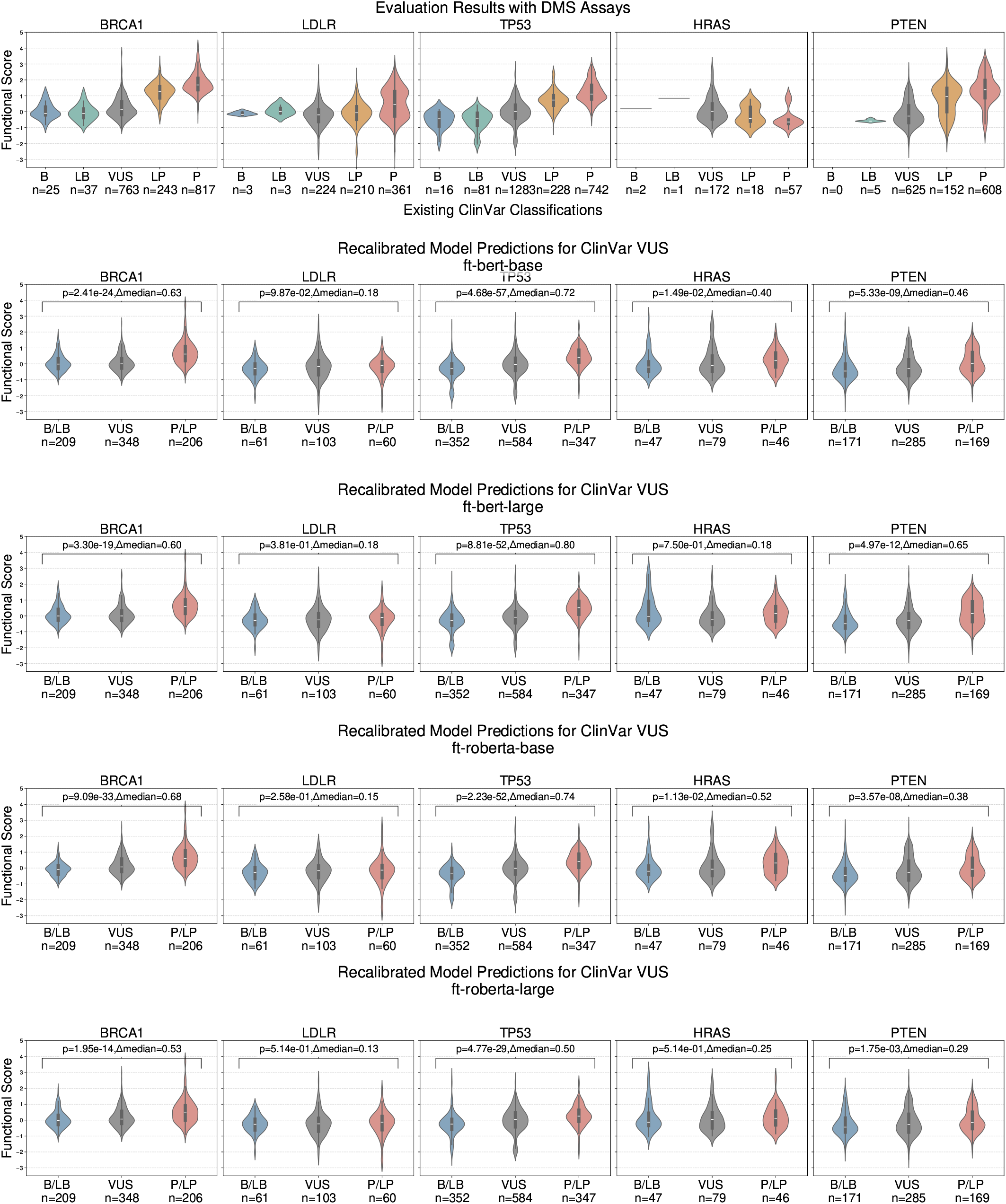
A comparison of multiple general-domain BERT-architecture models with different model sizes (BERT-base, BERT-large, RoBERTa-based, and RoBERTa-large) fine-tuned with **evidence-only** training data performances on DMS validation.

### DMS validation results with different thresholds

**Fig. 5:**
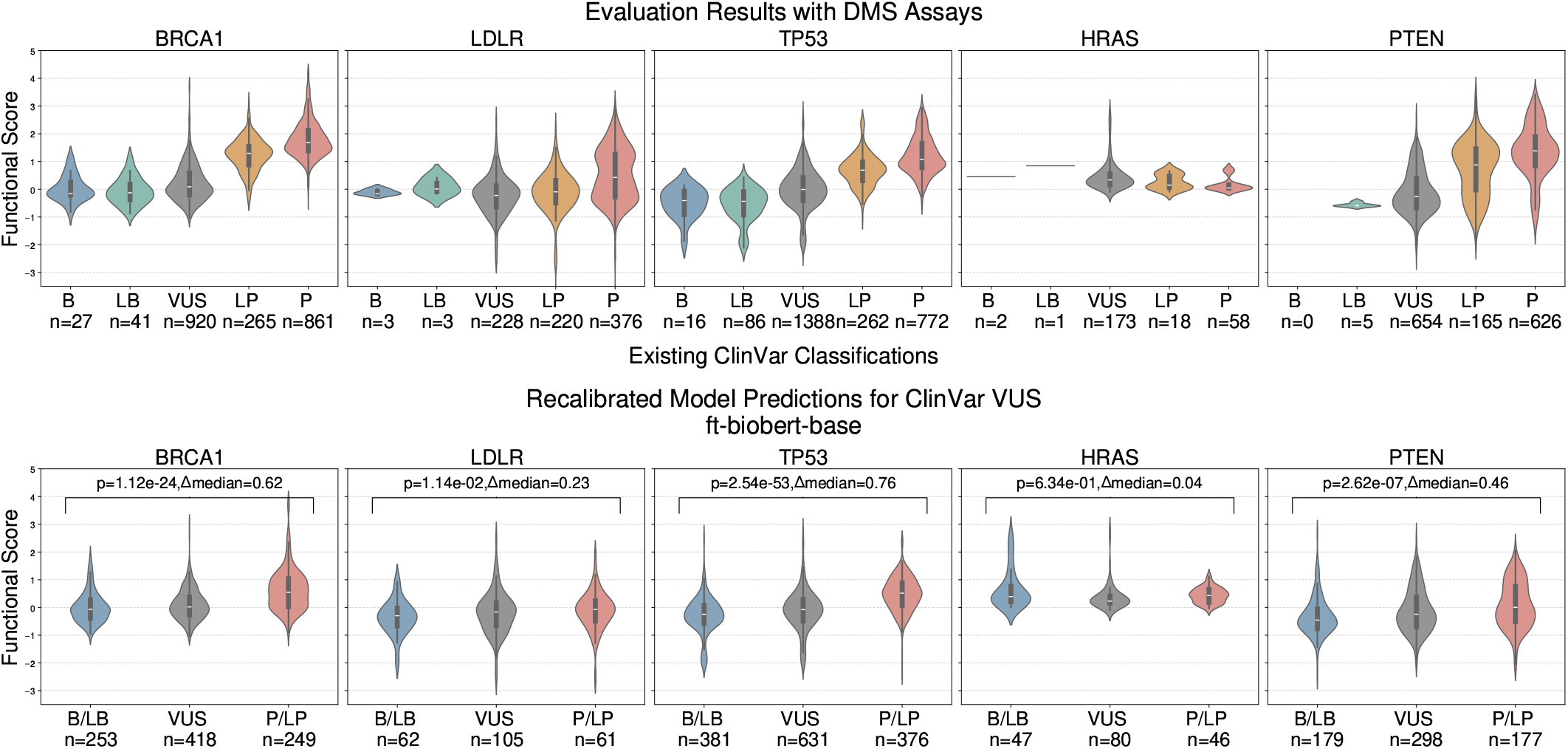
Top row: Existing classification on ClinVar (B, LB, VUS, LP, and P) on the *x*-axis and functional scores on the *y*-axis. Bottom row: recalibrated model prediction using 27.1% for P/LP prediction and 27.5% for B/LB prediction as the threshold on **VUS**-labeled submission on the *x*-axis (B/LB, VUS, and P/LP), and functional scores on the *y*-axis.

**Fig. 6:**
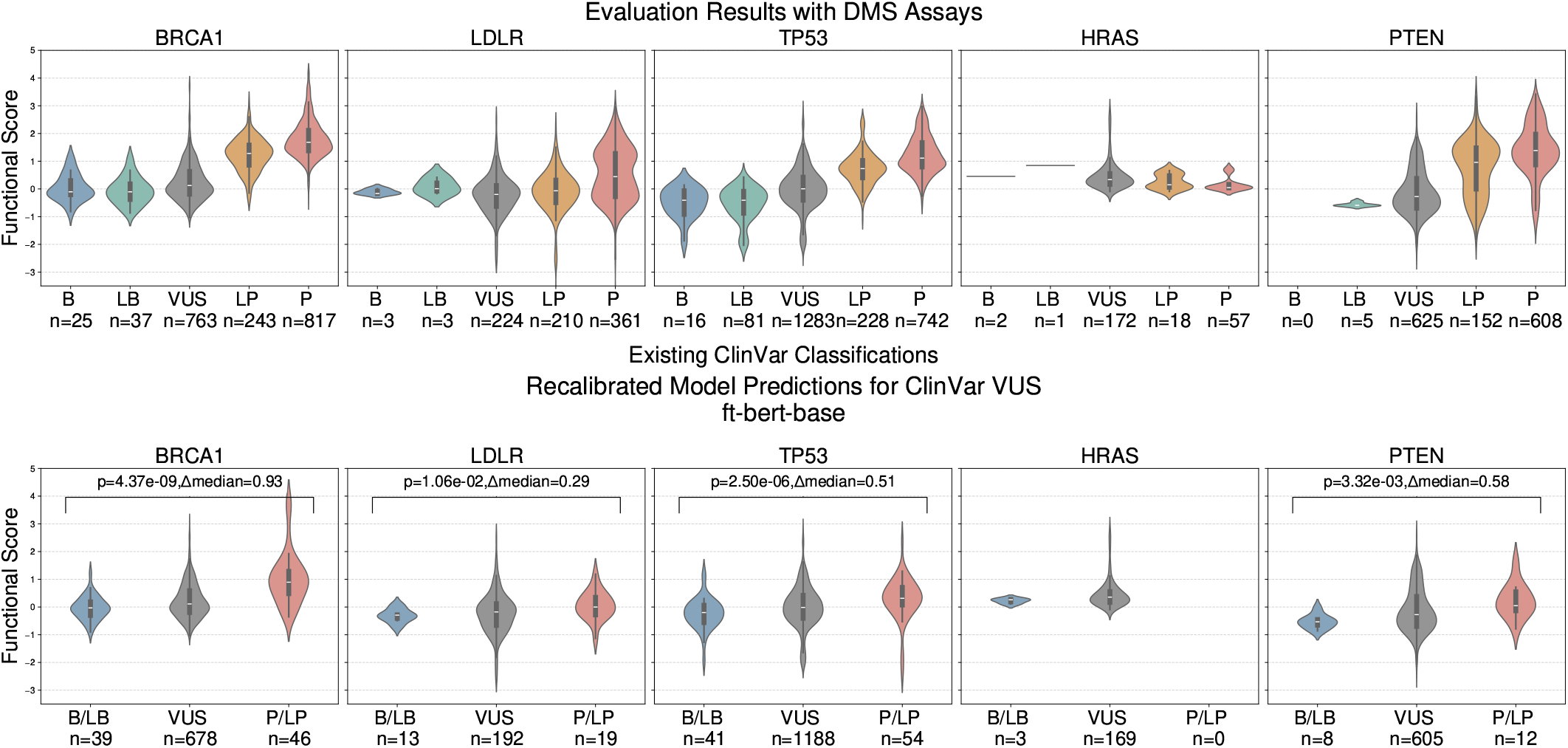
Top row: Existing classification on ClinVar (B, LB, VUS, LP, and P) on the *x*-axis and functional scores on the *y*-axis. Bottom row: directly plot model prediction of **VUS**-labeled submissions on the *x*-axis (B/LB, VUS, and P/LP) **without recalibration**, and functional scores on the *y*-axis

**Fig. 7:**
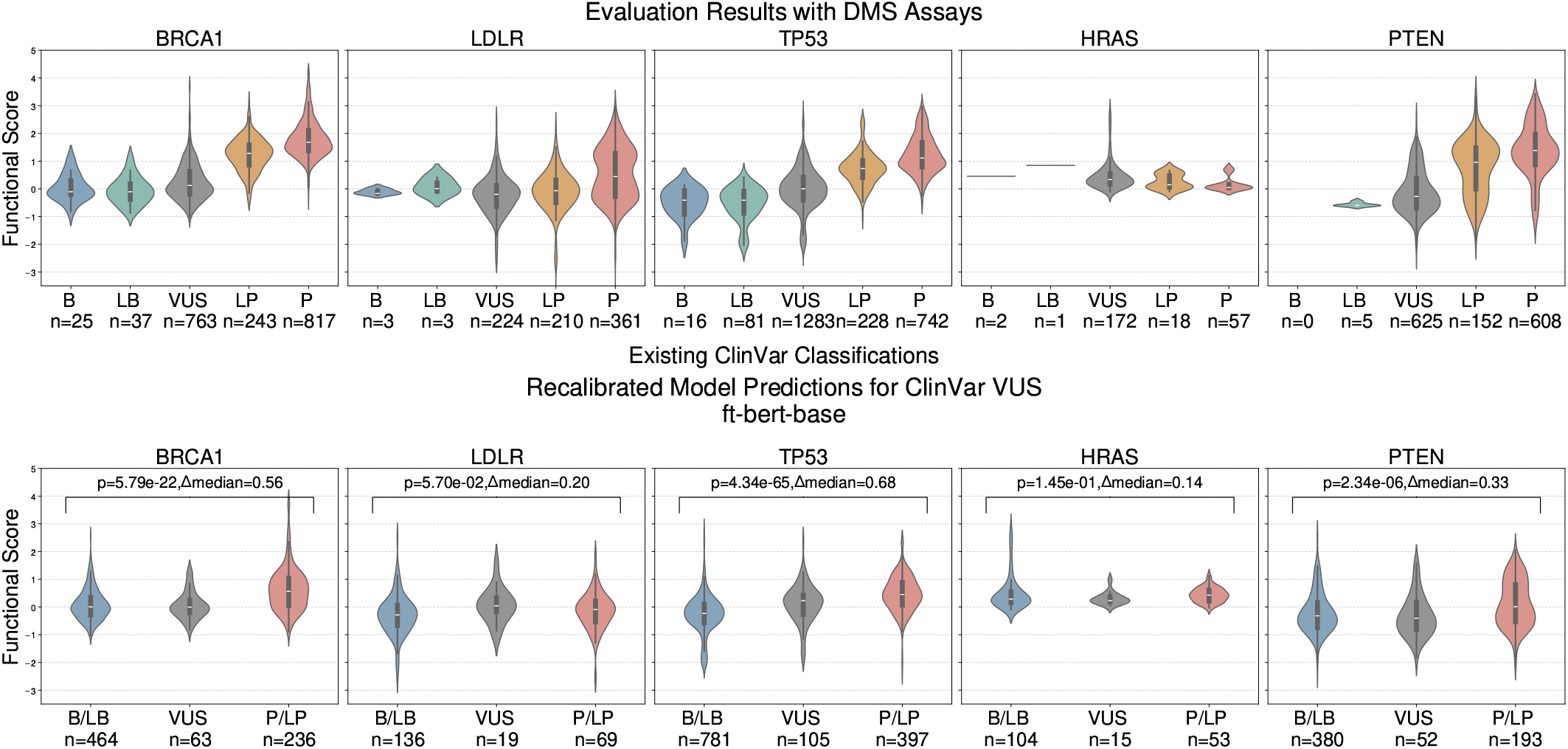
Top row: Existing classification on ClinVar (B, LB, VUS, LP, and P) on the *x*-axis and functional scores on the *y*-axis. Bottom row: recalibrated model prediction using 30.95% percentile for P/LP prediction and 60.91% percentile for B/LB prediction as the threshold (based on the distribution of VUS observed in 10,000 samples using the variant interpretation thresholds defined in AlphaMissense 36) on **VUS**-labeled submission on the *x*-axis (B/LB, VUS, and P/LP), and functional scores on the *y*-axis.

**Fig. 8:**
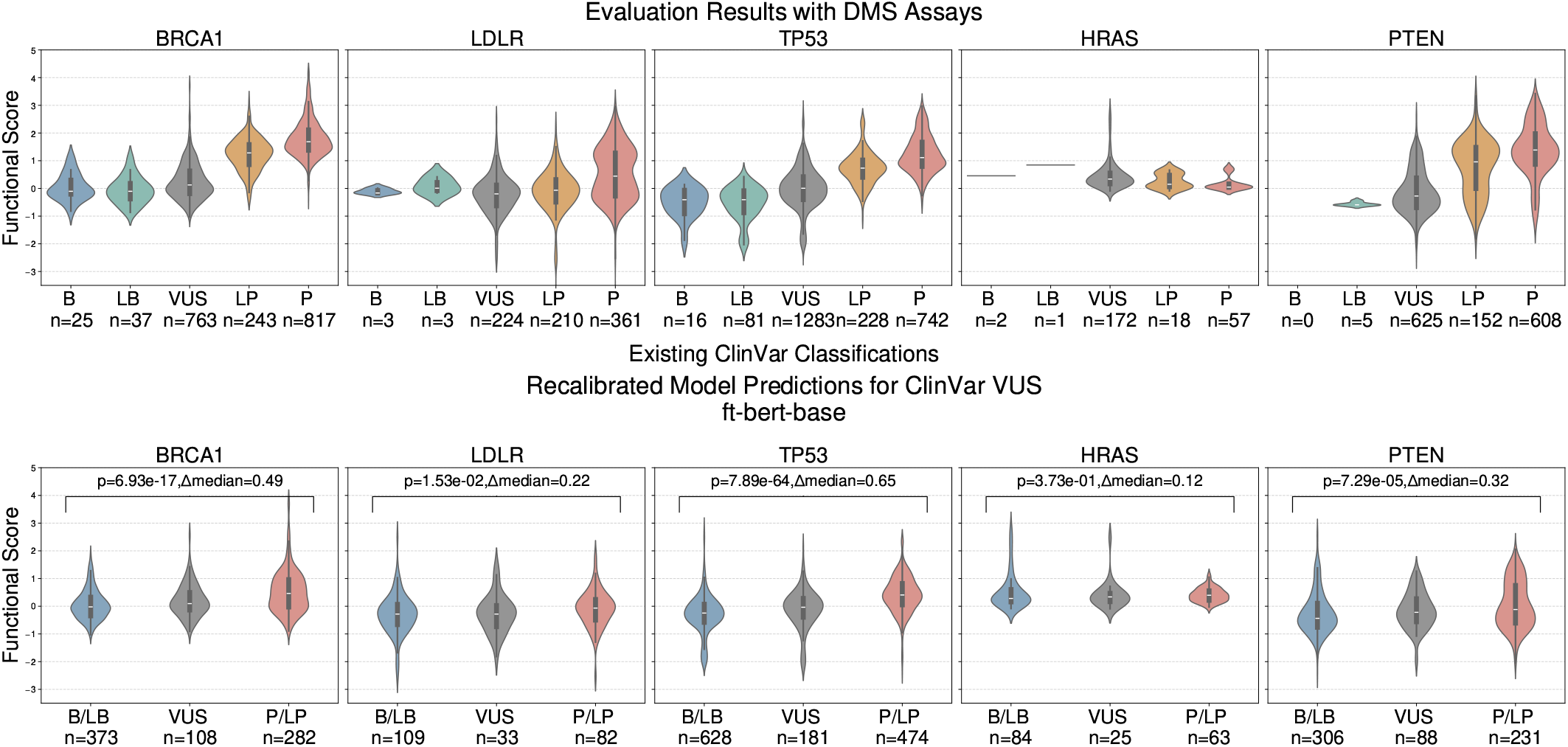
Top row: Existing classification on ClinVar (B, LB, VUS, LP, and P) on the *x*-axis and functional scores on the *y*-axis. Bottom row: recalibrated model prediction using 37% for P/LP prediction and 49% for B/LB prediction as the threshold (based on the distribution estimated from Figure 5 (B) in AlphaMissense 36) on **VUS**-labeled submission on the *x*-axis (B/LB, VUS, and P/LP), and functional scores on the *y*-axis.

### UMAP visualization clustering examples

#### Examples of VUS submissions closest to a P/LP cluster in UMAP visualization

**REVEL and AlphaMissense scores are not available for loss-of-function variants**

- **VCV:** VCV001768964

**SCV:** SCV002691514

**Comment:** The c.12860 12863delTTCA variant, located in coding exon 29 of the APOB gene, results from a deletion of 4 nucleotides at nucleotide positions 12860 to 12863, causing a translational frameshift with a predicted alternate stop codon (p.I4287Sfs*22). Frameshifts are typically deleterious in nature, however, this frameshift occurs at the 3’ terminus of APOB and is not expected to trigger nonsense-mediated mRNA decay. This alteration impacts the last 277 amino acids in the protein C-terminus, which has been indicated to play a role in both lipoprotein assembly and LDLR binding (McCormick SP et al. J. Biol. Chem., 1997 Sep;272:23616-22; Boren J et al. J. Clin. Invest., 1998 Mar;101:1084-93; Borén J et al. J. Biol. Chem., 2001 Mar;276:9214-8). Nevertheless, the exact functional impact of these altered amino acids is unknown at this time. Since supporting evidence is limited at this time, the clinical significance of this alteration remains unclear.

**gnomAD:** absent from gnomAD

**Variant type:** frameshift variant

- **VCV:** VCV000483568

**SCV:** SCV000669586

**Comment:** The p.Y97D variant (also known as c.289T¿G), located in coding exon 3 of the MLH1 gene, results from a T to G substitution at nucleotide position 289. The tyrosine at codon 97 is replaced by aspartic acid, an amino acid with highly dissimilar properties. This alteration is observed in an individual whose colorectal tumor demonstrated normal mismatch repair protein expression on immunohistochemistry (Ambry internal data). This alteration was reported in an individual whose colorectal tumor demonstrated low microsatellite instability and family history meeting Amsterdam criteria (Dominguez-Valentin M et al. Hered Cancer Clin Pract, 2013 Dec;11:18; Köger N et al. Genes Chromosomes Cancer, 2018 Jul;57:350-358). This alteration is also reported in an individual whose colorectal tumor demonstrated high microsatellite instability but weak MLH1 protein expression on immunohistochemistry, and family history meeting Bethesda criteria (Hardt K et al. Fam Cancer, 2011 Jun;10:273-84). Functional assays demonstrated reduced MMR activity for p.Y97D compared to wild-type MLH1; however, expression for p.Y97D was similar to wild-type MLH1 (Köger N et al. Genes Chromosomes Cancer, 2018 Jul;57:350-358). Based on internal structural analysis using published crystal structures, p.Y97D is moderately disruptive to the structure (Ambry internal data; (Ban C et al. Cell, 1999 Apr;97:85-97; Hu X et al. FEBS Lett, 2003 Jun;544:268-73; Wu H et al. Acta Crystallogr F Struct Biol Commun, 2015 Aug;71:981-5). This amino acid position is highly conserved in available vertebrate species. In addition, this alteration is predicted to be deleterious by in silico analysis. Since supporting evidence is conflicting at this time, the clinical significance of this alteration remains unclear.

**REVEL score:** 0.964

**AlphaMissense score:** 0.988

**gnomAD:** absent from gnomAD

**Variant type:** missense variant

**REVEL and AlphaMissense scores are not available for loss-of-function variants**

- **VCV:** VCV000066877

**SCV:** SCV002720118

**Comment:** The p.R654* variant (also known as c.1960C¿T), located in coding exon 11 of the LMNA gene, results from a C to T substitution at nucleotide position 1960. This changes the amino acid from an arginine to a stop codon within coding exon 11. This alteration occurs at the 3’ terminus of the LMNA gene, is not expected to trigger nonsense-mediated mRNA decay, and only impacts the last 11 amino acids of the protein. The exact functional effect of this alteration is unknown. This variant was reported in a family with dilated cardiomyopathy (DCM) with segregation in three affected family members, and absence in three other affected family members; an additional cardiac variant in PLN was also detected in the proband and the three affected family members without the LMNA variant (Parks SB et al. Am. Heart J., 2008 Jul;156:161-9; Cowan JR et al. Circ Genom Precis Med, 2018 Jul;11:e002038). This alteration was also detected in an individual with Hutchinson-Gilford Progeria syndrome, who was homozygous for a ZMPSTE24 loss of function variant, as well as in his unaffected mother and brother (Denecke J et al. Hum. Mutat., 2006 Jun;27:524-31). In addition, the variant was reported in an individual with Emery Dreifuss muscular dystrophy type 2 with symptoms of axonal neuropathy (Denecke J et al. Hum. Mutat., 2006 Jun;27:524-31; Bernasconi P et al. Nucleus, 2018 Jan;9:292-304). Functional studies showed formation of protein aggregates, but the functional impact of this finding has not been determined (Cowan J et al. Circ Cardiovasc Genet, 2010 Feb;3:6-14). Since supporting evidence is limited at this time, the clinical significance of this alteration remains unclear.

**gnomAD:** 2.464e-05

**Variant type:** stop gained

- **VCV:** VCV000546000

**SCV:** SCV001185259

**Comment:** The p.E17* variant (also known as c.49G¿T), located in coding exon 1 of the VHL gene, results from a G to T substitution at nucleotide position 49. This changes the amino acid from a glutamic acid to a stop codon within coding exon 1. Premature stop codons are typically deleterious in nature; however, an alternate initiation codon exists 37 amino acids downstream from this alteration, and is reported to result in a biologically active isoform known as VHL 19 (Iliopoulos O et al, Proc. Natl. Acad. Sci. U.S.A. 1998 Sep; 95(20):11661-6. Schoenfeld A et al, Proc. Natl. Acad. Sci. U.S.A. 1998 Jul; 95(15):8817-22). Since supporting evidence is limited at this time, the clinical significance of this alteration remains unclear.

**gnomAD:** 0

**Variant type:** stop gained

- **VCV:** VCV000426716

**SCV:** SCV000577240

**Comment:** The E227G variant has not been published as a pathogenic variant, nor has it been reported as a benign variant to our knowledge. The E227G variant is not observed in large population cohorts (Lek et al., 2016; 1000 Genomes Consortium et al., 2015; Exome Variant Server). This variant is a non-conservative amino acid substitution, which is likely to impact secondary protein structure as these residues differ in polarity, charge, size and/or other properties. This substitution occurs at a position that is conserved across species, and in silico analysis predicts this variant is probably damaging to the protein structure/function. However, missense variants in nearby residues have not been reported in the Human Gene Mutation Database in association with SEPN1-related disorders (Stenson et al., 2014).

**REVEL score:** 0.621

**gnomAD:** 6.841e-07

**Variant type:** missense variant

## Footnotes

1 https://www.ncbi.nlm.nih.gov/clinvar/

